# Characterizing the genetic architecture of Parkinson’s disease in Latinos

**DOI:** 10.1101/2020.11.09.20227124

**Authors:** Douglas Loesch, Andrea R. V. R. Horimoto, Karl Heilbron, Elif Irem Sarihan, Miguel Inca-Martinez, Emily Mason, Mario Cornejo-Olivas, Luis Torres, Pilar Mazzetti, Carlos Cosentino, Elison Sarapura-Castro, Andrea Rivera-Valdivia, Angel C. Medina, Elena Dieguez, Victor Raggio, Andres Lescano, Vitor Tumas, Vanderci Borges, Henrique B. Ferraz, Carlos R. Rieder, Artur Schumacher-Schuh, Bruno L. Santos-Lobato, Carlos Velez-Pardo, Marlene Jimenez-Del-Rio, Francisco Lopera, Sonia Moreno, Pedro Chana-Cuevas, William Fernandez, Gonzalo Arboleda, Humberto Arboleda, Carlos E. Arboleda-Bustos, Dora Yearout, Cyrus P. Zabetian, the 23andMe Research Team, Paul Cannon, Timothy A. Thornton, Timothy D. O’Connor, Ignacio F. Mata, on behalf of the Latin American Research Consortium on the Genetics of Parkinson’s Disease (LARGE-PD)

## Abstract

To date, over 90 Parkinson’s disease (PD) risk variants have been reported from genome-wide association studies (GWAS). However, these GWAS efforts have been limited to individuals of European and East Asian ancestry. We performed the first GWAS of Latino PD patients from South America, comparing 807 cases against 690 controls followed by association testing of suggestive loci in a replication cohort of 1,234 cases and 439,522 controls. We demonstrated that *SNCA* plays a significant role in PD etiology in a Latino cohort and identified a suggestive locus near *NRROS* on chromosome 3 that appeared to be driven by Peruvian subjects. We also characterized the overlap of PD genetic architecture between Europeans and Latinos with a replication of significant variants identified by Nalls et al. in their 2019 GWAS^1^, finding 80% concordance in direction of effect. We then leveraged the population history of Latinos via admixture mapping, identifying a significant locus on chromosome 14 in a joint test of ancestries, driven by the Native American ancestral background, and a significant locus on chromosome 6 in our test of African ancestry, containing the genes *STXBP6* and *RPS6KA2*, respectively. Ultimately, our work reflects the most comprehensive characterization of PD genetic architecture in Latinos to date.

## Introduction

Parkinson’s disease (PD) is the second most common neurodegenerative disorder after Alzheimer’s disease and as the fastest growing neurological disorder is expected to impose an increasing social and economic burden worldwide.^1,2^ PD prevalence increases with age, reaching 1-2% in people over age 65.^2,3^ Though age remains the largest single risk factor for PD, specific environmental exposures and genetics also play a role in PD etiology. ^1,2,4^ While rare genetic variants have been shown to play a role in familial forms of the disease, the majority of patients do not report any family history.^5^ In these individuals, candidate gene approaches and large-scale genome-wide association studies (GWAS) have identified common genetic variants which contribute to PD risk.^3^

PD is truly a global disease, impacting all ethnic groups. Despite this, GWAS efforts to date have been limited to individuals of European and East Asian ancestry.^1,6–9^ This under-representation is not limited to PD; nearly 80% of all study participants represented in the GWAS Catalog are of European descent.^10,11^ This lack of diversity is even more evident when further broken down by ancestry; as of 2018, only 1.3% of study participants in the GWAS Catalog are Hispanics/Latinos, 0.03% are Native American, and 2.4% are African.^11^ This risks missing out on population-specific variation, and creating biased polygenic risk scores due to linkage disequilibrium structure specific to the European subjects used to generate the majority of GWAS summary statistics.^11–13^

PD incidence rates are rising in nearly every global region^2^, highlighting the need for greater diversity in PD consortiums. A study of Medicare beneficiaries and a study of Kaiser Permanente members both found the age-adjusted PD incidence rate to be highest in Hispanics/Latinos among the surveyed ancestries.^14,15^ Furthermore, few genetic studies have been done in Latin America and the existing studies have exclusively utilized candidate gene approaches.^16–20^ The Latin American Research Consortium on the Genetics of PD (LARGE-PD) was formed in 2009 to fill this gap.^21^ LARGE-PD is an ongoing effort that includes 35 institutions in 12 countries across the Americas and the Caribbean. Here we performed the first GWAS of Latino PD patients from South America composed of 1,497 subjects from LARGE-PD and 8.7 million variants obtained using a genotyping array and an imputation reference panel optimized for diverse subjects.^22,23^

## Methods

### Sample Description

1,504 LARGE-PD samples from Uruguay, Peru, Chile, Brazil, and Colombia using the Multi-Ethnic Genotyping Array (MEGA) from Illumina^22^ were genotyped at the Genomics Core at the University of Washington. After performing standard quality control steps (described below), we selected 807 PD cases (mean age of 61.7 years and 53% males) and 690 controls (mean age of 56.5 years and 33% males) (**see Supplementary Table 1 for the complete cohort description)**. PD patients were evaluated by a local movement disorder specialist using the UK PD Society Brain Bank clinical diagnostic criteria (UKPDSBB).^24^ Individuals who did not exhibit neurological symptoms were selected as controls. All participants provided written informed consent according to their respective locale’s national requirements.

### Genome-Wide Association Analysis (GWAS)

#### Quality Control

We converted the raw genotype data to PLINK format and carried out quality control (QC) steps using PLINK 1.9.^25^ We removed unplaced, duplicated, non-autosomal, monomorphic variants prior to filtering. We also filtered for HWE using a p-value threshold of less than 1×10^−06^ in controls and 1×10^−10^ in cases^26^ and a genotype missingness filter of 5%. No samples failed due to missing greater than 5% of genotyped sites and the ascertained sex of all samples matched the sex inferred from the X chromosome. We flagged three pairs of samples as either duplicates or monozygotic twins via PLINK’s identity-by-descent procedure; these samples are likely the same individual. For this study, we excluded all six samples. In addition, we excluded one individual whose diagnosis had changed in the face of new clinical data. Overall, 1,497 samples and 1,240,909 bi-allelic variants passed QC with an overall genotyping rate of 0.999.

#### Imputation

We imputed the LARGE-PD dataset using the TOPMed Imputation Server (version r1) which utilizes MINIMAC4 and a reference panel of 125,568 haplotypes from diverse samples.^27^ Variants unable to be lifted over to hg38 or rectified via strand flips were removed by the Imputation Server pipeline. We retained imputed variants if they had a minimum imputation R^2^ greater than 0.3. For analyses requiring hg19 coordinates, we lifted the imputed results back to hg19 using Picard Tools.^28^

#### Characterization of LARGE-PD Population Structure

To improve inference of LARGE-PD population structure, we merged LARGE-PD genotyped variants with sequenced variants from the 1000 Genomes Project^29^; the intersection consisted of 606,977 variants. We then filtered the merged dataset for a minimum minor allele frequency (MAF) of 0.01 and linkage disequilibrium (LD) pruning using PLINK’s indep-pairwise with a window of 50 variants, a step of 5 variants and a maximum R^2^ of 0.2 as its parameters. For the admixture analysis, we resolved pairs of relatives by randomly removing one relative from each pair using KING’s unrelated algorithm^30^ and a threshold of second-degree relatedness. We ran ADMIXTURE^31^ with K equal to 3, 4, or 5. For the K=5, we included all 1000 Genomes populations; for K =4, we removed South Asian samples; for K=3, we removed East Asian and South Asian samples. We repeated each analysis 20 times using the random seed option and retained the repetition with the highest log-likelihood. In addition to the admixture analysis, we performed principal component analysis (PCA) on all LARGE-PD subjects using the PC-AiR^32^ and PC-Relate^33^ methods that are implemented in the GENESIS package and is available from Bioconductor^34^ (**see supplementary methods**).

#### Estimation of Additive Heritability (h^2^)

We estimated heritability using GCTA^35,36^ and imputed LARGE-PD variants and a method developed by Yang et al. to correct for the bias due to LD.^37^ Imputed variants with a MAF of at least 1% are stratified into four groups based on their LD score, followed by the estimation of genetic relatedness matrices (GRMs) corresponding to each of the strata. We restricted our heritability analysis to the unrelated subset of LARGE-PD up to the second degree, as determined via KING^30^ in the same manner described in the admixture analysis. We then estimated narrow-sense heritability using AI-REML in GCTA^36^ and the four stratified GRMs, assuming a prevalence of 0.5% and including age, sex, the first five PCs, and recruitment site as fixed effects.

#### Genome-Wide Association Study

We conducted a GWAS utilizing all samples from the imputed LARGE-PD cohort and logistic mixed models implemented in the GENESIS R package.^38^ We included age, sex, the first five PCs, and the GRM estimated using GCTA^36^ in our null model. We tested imputed dosages against the null via a score test.

#### Fine Mapping

We assessed the regional association plots prepared using the LocusZoom tool^39^, identified the variants previously associated with PD in the GWAS Catalog^40^, and obtained additional functional annotations using the Ensembl Variant Effect Predictor.^41^ We determined the LD structure of the chromosome 4 peak using PLINK 1.9. We also utilized this LD information to create custom LocusZoom-style plots. We determined the 95% credible set using PAINTOR 3.0^42^ (**see supplementary methods**).

#### Conditional Analysis

We performed a conditional analysis where we adjusted for rs356182, the lead *SNCA* variant in European-ancestry PD analyses along with age, sex, and the first 5 PCs using logistic mixed models implemented with the GMMAT package^43^ in R. We evaluated p-values using two different p-value thresholds: the number of GWAS-significant variants and the number of independent tests in the *SNCA* region.^44^ We then performed a stepwise conditional analysis, adjusting for rs356182 and additional significant SNPs until no SNPs remained statistically significant.

#### 23andMe Replication of LARGE-PD GWAS Primary Results

We selected 180 variants for replication with a minimum p-value of 1×10^−5^ provided they met one of the following criteria: the top variant at a genomic locus (+/- 500 KB) or in the 95% credible set at the *NRROS* and *SNCA* loci. 23andMe tested the set of identified variants via their replication pipeline and an independent cohort of 1,234 Hispanic/Latino subjects with self-reported PD status and 439,522 controls. All self-reported PD cases and controls from 23andMe provided informed consent and answered surveys online according to 23andMe’s protocol, which was reviewed and approved by Ethical & Independent Review Services, a private institutional review board (http://www.eandireview.com). Samples were genotyped on one of five genotyping platforms; for inclusion, samples needed a minimal call rate of 98.5%. Genotyped samples were then phased using either Finch or Eagle2^45^ and imputed using Minimac3 and a reference panel of 1000 Genomes Phase III^29^ and UK10K data.^46^ For this replication study, samples were classified as Latino using a genotype-based pipeline^47^ consisting of a support vector machine and a hidden Markov model, followed by a logistic classifier to differentiate Latinos from African-Americans. Unrelated individuals were included in the analysis, as determined via identity-by-descent (IBD). Variants were tested for association with PD status using logistic regression, adjusting for age, sex, the first five PCs, and genotyping platform.Reported p-values were from a likelihood ratio test (see **supplementary methods**).

#### Replication of Previously Identified PD Risk Variants

We attempted to test 90 independent PD risk variants, previously identified by Nalls et al. 2019^1^, in LARGE-PD for association with PD. We successfully imputed 84 of the 90 variants. 5 of the six variants that we were unable to impute were absent from the TOPMed imputation reference panel due to failing TOPMed’s QC protocol; the remaining variant was absent from the dataset. For this variant look-up, we applied the approximation of the Wald test to the score test results from our primary GWAS in order to obtain beta coefficients. In order to ensure fair comparisons, we removed strand ambiguous (CG/AT) sites with a MAF greater than 0.30. We also removed rare variants with a minor allele count (MAC) of less than or equal to 10 in LARGE-PD. Beta coefficient correlations were performed using Pearson’s method. In addition to the variants from Nalls et al. 2019^1^, we also performed a variant look-up of additional PD GWAS results from European and East Asian-ancestry studies.^6,7,9^

### Admixture Mapping

#### Quality Control

For the admixture mapping, we employed a slightly modified quality control pipeline. We converted the Illumina files to binary PLINK^48^ format. We excluded SNPs with missing genotype > 0.10, HWE p-value <.0001, and monomorphic SNPs, with a final genotyping rate of 0.998. We did not need to exclude any of the subjects for low genotyping (maximum missing genotype data of 0.10). The final admixture mapping analysis included all 1,497 subjects with both genotype and phenotype data, and 1,294,079 SNPs that passed quality control filtering.

#### Admixture Mapping Analysis

We selected 63 unrelated individuals from CEU (Utah residents with Northern and Western European ancestry from the CEPH collection) and YRI (Yoruba in Ibadan, Nigeria) samples from the HapMap project phase III^49^ (International HapMap Consortium, 2003), and Native American (Pima, Maya and Colombian) samples from the HGDP project (https://www.hagsc.org/hgdp/) to be used as references for European, African, and Native American ancestral populations. We excluded 242 CEU and YRI samples from the dataset in order to keep balanced reference samples (63 samples for each ancestral reference population), as recommended in the RFMix manual.^50^ We then merged the HapMap and HGDP reference datasets with our 1,497 LARGE-PD samples using PLINK, keeping 164,651 autosomal SNPs in common to all datasets with an overall genotyping rate of 0.999. We performed a joint phasing of LARGE-PD and reference samples using Shapeit2^51^ and an additional reference panel of phased haplotypes from 1000 Genomes project, phase III.^29^

We performed the local ancestry estimation using RFMix^50^, version 1.5.4, considering the trihybrid ancestry (European, African, and Native American) of the samples. We prepared the input files for RFMix using auxiliary Python scripts of the Ancestry Pipeline developed by Martin et al. 2017.^13^

We performed admixture mapping through a joint test implemented in the GENESIS R package^34^ (https://github.com/UW-GAC/GENESIS), in which all European, African, and Native American ancestries are tested jointly in an admixture mapping logistic mixed model. The analysis was performed in two steps. Firstly, we fit the logistic mixed model under the null hypothesis of no genetic effect including sex, age, and the first five components as fixed effects and the genetic relationship matrix (**see supplementary methods)** as random effects. Then, this fitted null model was used in a second step in which we conducted a multivariate score test to verify the association between the ancestry at each locus and PD status (**see supplementary methods** for a detailed description of the admixture mapping model).

Secondary admixture mapping analyses were performed for each European, African, and Native American ancestry separately in order to identify which ancestral population was driving the significant signal. Based on previous studies, a p-value of 5×10^−5^ controls the type I error at level of 0.05^52^ (**see supplementary methods**). We fine-mapped the suggestive admixture peaks by overlaying our GWAS results (as described above) with admixture mapping peaks. Significance levels were determined via Bonferroni’s correction for the number of imputed SNPs with minimum MAF of 0.01 in each peak.

## Results

### Cohort Description and Ancestry Analysis

We genotyped LARGE-PD samples using the Illumina Multi-Ethnic Genotyping Array (MEGA) which was designed to accurately genotype diverse samples and provides suitable coverage for imputation. The samples came from PD cases and healthy controls across nine sites in five countries: Uruguay, Brazil, Colombia, Peru, and Chile (**see supplementary table 1**). LARGE-PD cases were 53% male and had a mean age of 61.7 years (+-12.8 years) and a mean age at onset of 54.1 years (+- 14.4 years); controls were 33% male and had a mean age of 56.5 years (+- 14.6 years). Hispanic/Latino populations tend to have a three-way admixture pattern with contributions from African, European, and Native American ancestry. The exact proportions of these ancestries can vary dramatically and typically reflect the demographic history of the region. Restricting LARGE-PD to unrelated subjects, the mean proportion of African ancestry was 0.0517, Amerindian ancestry was 0.47, European ancestry was 0.47, and other ancestries were 0.0076 (**see Figure 1; supplementary table 2)**. The mean proportion of Amerindian ancestry was highest in Peru-Puno (0.99) and lowest in Brazil-Ribeirao Preto (0.063). African ancestry was highest in Brazil-Sao Paulo (0.14) and lowest in Peru-Puno (1×10^−05^). European ancestry was highest in Uruguay (0.825) and lowest in Peru-Puno (0.007). Note that migration was not limited to these three populations; East Asian ancestry was observed in several individuals (**see supplementary figure 1**).

**Figure 1:**
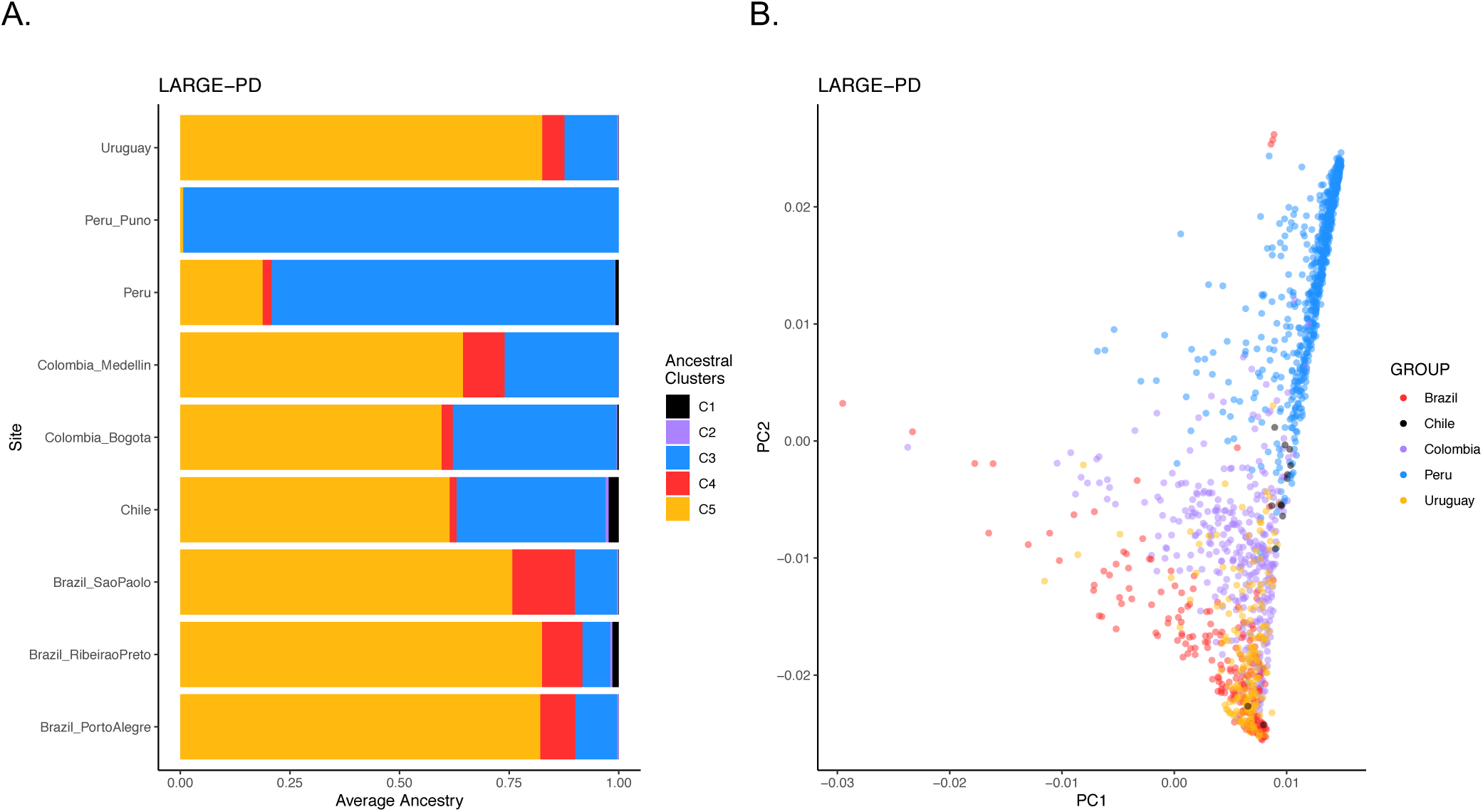
LARGE-PD Demography. *1A: Mean ancestry proportions by LARGE-PD site*. We estimated ancestry proportions using ADMIXTURE and a K of 5 in a joint dataset that included LARGE-PD and 1000 Genomes Project samples.^74^ Using 1000 Genomes super-population codes to infer the ancestry underlying each cluster, C1 represents East Asian, C2 represents South Asian, C3 represents Native American, C4 represents African, and C5 represents European ancestry (see Figure S1). *1B: PCA plot of LARGE-PD subjects*. We conducted a principal components analysis using PC-AiR in the merged 1000 Genomes-LARGE-PD dataset. Note the preponderance of individuals with high Amerindian and European ancestries. Principal components were calculated using the PC-AiR algorithm from the GENESIS package in R.

### Additive Heritability of PD

Using GCTA and all imputed SNPs with a minor allele frequency (MAF) of at least 0.01, we estimated the additive heritability (h^2^) of PD in LARGE-PD to be 0.38 (SE 0.068) with a prevalence of 0.5%. We used a method that accounts for the bias in sequence and imputed data caused by LD (linkage disequilibrium; **see methods**).

### Genome-wide Association Study

We imputed the genotyped data with the TOPMed imputation server, after extensive quality control steps (see methods). This imputation server has been shown to improve imputation for Hispanics/Latinos.^23,27^ We tested variants with a minimum MAF of 1% for association with the disease using a logistic mixed model as implemented by the GENESIS package (see methods). One locus achieved genome-wide significance: the *SNCA* locus on chromosome 4 (**see Table 1; Figure 2**). At this locus, rs356225 achieved the lowest p-value (4.22×10^−9^). A second locus in chromosome 3 appears suggestive, with rs78820950 achieving the lowest p-value (8.25 ×10^−8^). This locus is located in an intergenic region between *FBXO45* and *NRROS*. Overall, we observed minimal inflation (GC lambda 1.017); consequently, we did correct for this inflation factor (**see supplementary figure 2**).

**Table 1:** LARGE-PD GWAS-significant results.

| SNP | CHROM | POS | FREQ | P.VALUE | REF | ALT | REPLICATES | GENE | R2 | R2_PERU | R2_EUR |
| --- | --- | --- | --- | --- | --- | --- | --- | --- | --- | --- | --- |
| rs356183 | 4 | 89704947 | 0.492 | 6.10E-09 | G | C | NA | SNCA | 0.217 | 0.419 | 0.641 |
| rs356182 | 4 | 89704960 | 0.556 | 2.48E-08 | G | A | YES | SNCA | NA | NA | NA |
| rs356181 | 4 | 89704988 | 0.481 | 2.60E-08 | G | A | NA | SNCA | 0.689 | 0.840 | 0.589 |
| rs356211 | 4 | 89715267 | 0.463 | 4.37E-08 | C | T | YES | SNCA | 0.612 | 0.802 | 0.492 |
| rs356219 | 4 | 89716450 | 0.522 | 4.50E-08 | G | A | YES | SNCA | 0.749 | 0.873 | 0.770 |
| rs356220 | 4 | 89720189 | 0.52 | 3.00E-08 | T | C | YES | SNCA | 0.744 | 0.870 | 0.763 |
| rs356221 | 4 | 89721313 | 0.47 | 6.15E-09 | A | T | YES | SNCA | 0.628 | 0.806 | 0.516 |
| rs356223 | 4 | 89722356 | 0.471 | 4.63E-09 | A | G | YES | SNCA | 0.631 | 0.806 | 0.517 |
| rs356225 | 4 | 89722606 | 0.47 | 4.22E-09 | C | G | YES | SNCA | 0.629 | 0.806 | 0.516 |
| rs356165 | 4 | 89725735 | 0.515 | 1.35E-08 | G | A | YES | SNCA | 0.742 | 0.870 | 0.767 |
| rs356204 | 4 | 89742391 | 0.47 | 5.63E-09 | T | C | YES | SNCA | 0.625 | 0.800 | 0.517 |
| rs356203 | 4 | 89744890 | 0.516 | 2.15E-08 | C | T | YES | SNCA | 0.741 | 0.865 | 0.767 |
| rs356200 | 4 | 89747463 | 0.47 | 5.77E-09 | T | C | YES | SNCA | 0.625 | 0.800 | 0.516 |
| rs189596 | 4 | 89750185 | 0.47 | 4.72E-09 | G | A | YES | SNCA | 0.626 | 0.802 | 0.516 |
| rs356168 | 4 | 89753280 | 0.47 | 4.69E-09 | G | A | YES | SNCA | 0.626 | 0.802 | 0.516 |
| rs2736990 | 4 | 89757390 | 0.474 | 5.85E-09 | G | A | YES | SNCA | 0.626 | 0.802 | 0.519 |
| rs356198 | 4 | 89761353 | 0.255 | 4.05E-08 | C | T | NOMINAL | SNCA | 0.217 | 0.419 | 0.020 |
| rs356197 | 4 | 89761599 | 0.255 | 4.05E-08 | G | A | NOMINAL | SNCA | 0.217 | 0.419 | 0.020 |
| rs356196 | 4 | 89761652 | 0.255 | 4.05E-08 | A | T | NA | SNCA | 0.217 | 0.419 | 0.020 |
| rs356191 | 4 | 89766969 | 0.272 | 2.76E-08 | G | A | NOMINAL | SNCA | 0.230 | 0.426 | 0.024 |
| rs356162 | 4 | 89776006 | 0.273 | 3.55E-08 | T | C | NOMINAL | SNCA | 0.228 | 0.426 | 0.024 |
| rs184810 | 4 | 89776828 | 0.273 | 3.54E-08 | T | C | NOMINAL | SNCA | 0.228 | 0.426 | 0.024 |
| rs3775434 | 4 | 89781630 | 0.32 | 1.15E-08 | A | G | YES | SNCA | 0.298 | 0.553 | 0.005 |
| rs3822089 | 4 | 89782860 | 0.319 | 1.31E-08 | G | A | YES | SNCA | 0.299 | 0.556 | 0.005 |
| rs3822090 | 4 | 89783725 | 0.319 | 1.29E-08 | C | T | YES | SNCA | 0.299 | 0.556 | 0.005 |
| rs3775439 | 4 | 89788590 | 0.321 | 2.02E-08 | G | A | YES | SNCA | 0.295 | 0.546 | 0.004 |
| rs2737029 | 4 | 89790619 | 0.484 | 2.11E-08 | T | C | YES | SNCA | 0.621 | 0.776 | 0.434 |
| rs6830166 | 4 | 89823842 | 0.326 | 9.35E-09 | C | T | YES | SNCA | 0.293 | 0.548 | NA |

LARGE-PD GWAS-significant results. SNP, the rs ID of the variant. CHROM, the chromosome. POS, the position in build HG38 coordinates. FREQ, the allele frequency observed in LARGE-PD. P.VALUE, the p-value from a score test. REF and ALT, the reference and alternative alleles. REPLICATES, indicates whether the variant replicated in the 23andMe cohort. GENE, the nearest gene. R2, the measure of LD (in $R^2$ ) of the variant with rs356182, the lead variant in European cohorts. R2\_PERU, the measure of LD with rs356182 in the Peruvian subset of LARGE-PD. R2\_EUR, the measure of LD with rs356182 in 1000 Genomes European populations.

**Figure 2:**
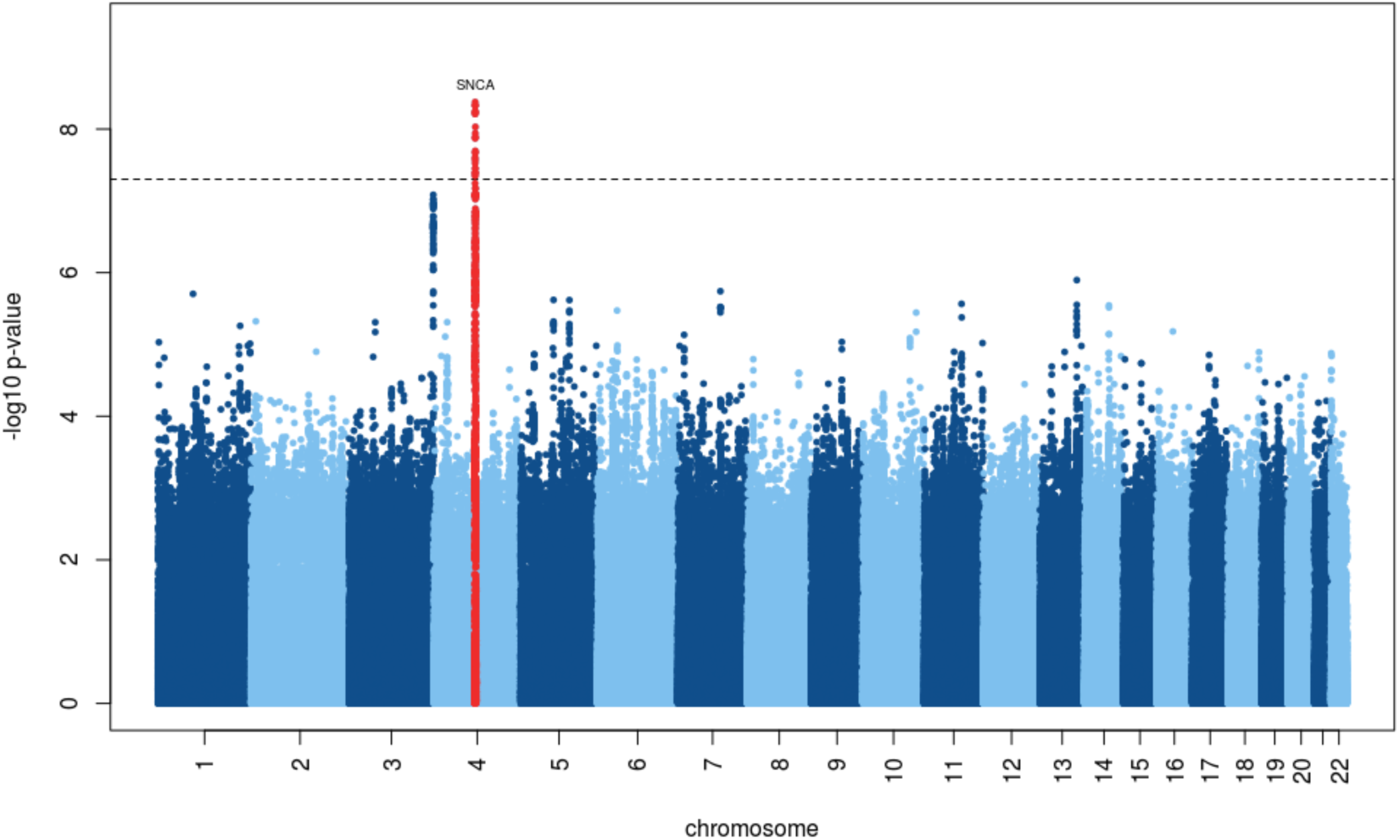
LARGE-PD GWAS results. Manhattan plot of log-transformed p-values by chromosome. P-values were obtained via a logistic mixed model adjusting for age, sex, and the first five principal components using the GENESIS package in R. The significant peak is located within *SNCA* on chromosome 4. The suggestive peak one near chromosome 3 is near *NRROS*.

The chromosome 3 locus has not been previously reported in the PD literature and is located in an intergenic region where the closest gene is *NRROS. NRROS* does appear to have a neurological function^53^, but PD-related evidence is limited. The most significant variant at this locus, rs78820950, has a MAF of 0.103 in LARGE-PD. However, this variant was more than three times as frequent in Peru than other LARGE-PD sites (0.168 vs. 0.045).

The chromosome 4 locus is well-characterized in PD literature and a number of SNPs have been put forth as contributing to PD risk.^1,44,54,55^ In LARGE-PD, 28 *SNCA* SNPs achieved genome-wide significance (**Table 1)**. By utilizing LD information, we observed three LD blocks **(see Figure 3**). Two of the three blocks contain well-documented PD SNPs^44,55–58^; the third contained a SNP associated with PD in one study.^59^ The top SNP, rs356225, is in strong LD with several known PD SNPs, with an R^2^ of 0.63 with rs356182 (**supplementary figure 3**). An overall pattern of higher LD was observed in the Peruvian subset (**supplementary figure 4)** than in the entire LARGE-PD cohort **(Figure 3)**.

**Figure 3:**
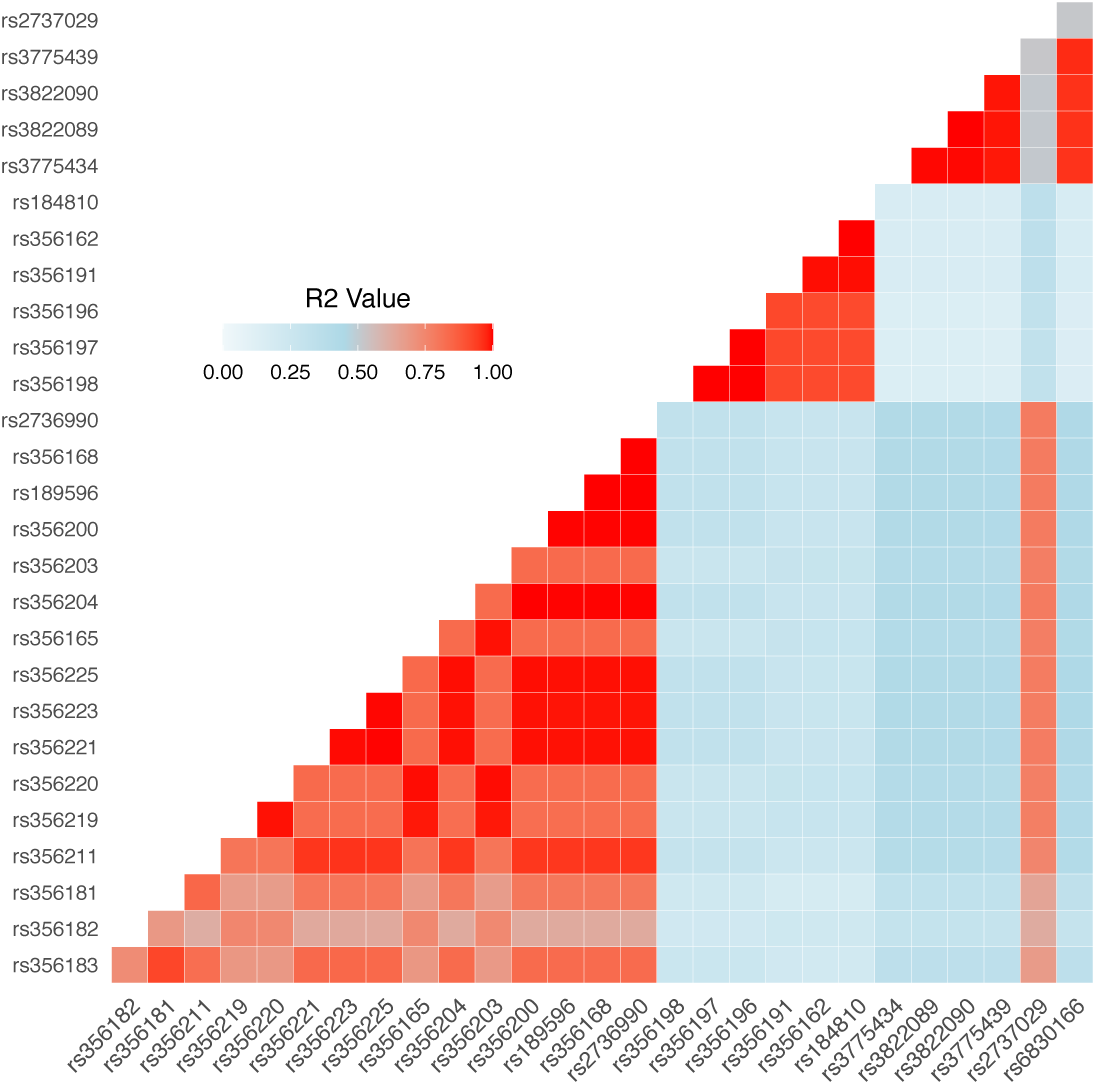
LD disequilibrium structure of GWAS significant SNCA variants. R^2^ values between each variant were obtained using PLINK 1.9 and are displayed as a correlation matrix; red indicates higher R^2^ values. Three independent LD blocks were observed with R^2^ values less than 0.5 between each block.

### Conditional Analysis

Using a logistic mixed model, we performed a conditional analysis where we adjusted for rs356182, the lead variant in European-ancestry PD meta-analyses, to test if it was driving the signal at this locus (**supplementary figure 5)**. When correcting for the number of GWAS-significant variants, 8 SNPs remain significant, though attenuated, after adjusting for rs356182, with rs6830166 having the smallest adjusted p-value (0.012). None of the SNPs remain statistically significant when adjusting for both rs356182 and rs6830166 (**see supplementary table 3)**, despite LD patterns showing evidence of three blocks. However, if we utilized a more stringent threshold, such as the regional correction implemented by Pihlstrøm et al. (n=220) in their conditional analysis of *SNCA*^44^, then we found minimal evidence of independence from rs356182 (**see supplementary table 3)**. A more comprehensive conditional analysis is necessary to pinpoint the number of independent *SNCA* signals implicated in Latino PD etiology.

### 23andMe Replication

We employed a relaxed criterion (p-values <1×10^−5^) to identify SNPs for replication (**see methods**). 23andMe tested each SNP using their replication pipeline (**see methods**) and a cohort of 1,234 self-reported PD cases and 439,522 controls. Self-reported ancestry was not utilized; rather, Latino ancestry was determined from the subject genotypes (**see methods)**. SNPs that were not directly genotyped were imputed. Each SNP was tested using logistic regression, adjusting for age, sex, the first 5 PCs, and genotype platform. Only the chromosome 4 locus replicated, with rs356182 achieving genome-wide significance (**see Table 1; supplementary table 4**), confirming the importance of *SNCA* and the variant rs356182 in particular.

### Replication of Known PD Loci

The largest PD-GWAS meta-analysis to date identified 90 independent GWAS-significant PD risk variants in subjects of European ancestry.^1^ To determine whether these SNPs conferred risk in the LARGE-PD cohort, we looked up 84 of the 90 SNPs in our primary GWAS (**supplementary Table 5)**. 76 of these variants passed our frequency and CG/AT filters (**see methods**). Sixty-three of the 76 variants (82.9%) had concordant direction of effect with a Pearson’s correlation of 0.82 (p < 2×10^−16^; **figure 4**). 10 variants were nominally significant (p < 0.05 and > 5.95×10^−04^), and two were significant after correction for 84 tests (rs356182 at the *SNCA* locus and rs117615688 close to *CRHR1*, p < 5.95×10^−04^). The set of variants with a MAF less than 0.0452 contains every variant with a difference in LARGE-PD and Nalls et al. beta coefficients greater than one standard deviation from the mean (**supplementary figure 6)**. This is likely due to a combination of the tendency of rare variants to have larger effect sizes and inaccuracies in the estimates of their beta coefficients. If we remove all variants with a MAF less than 0.05, the concordance rate improves to 86.3%.

**Figure 4:**
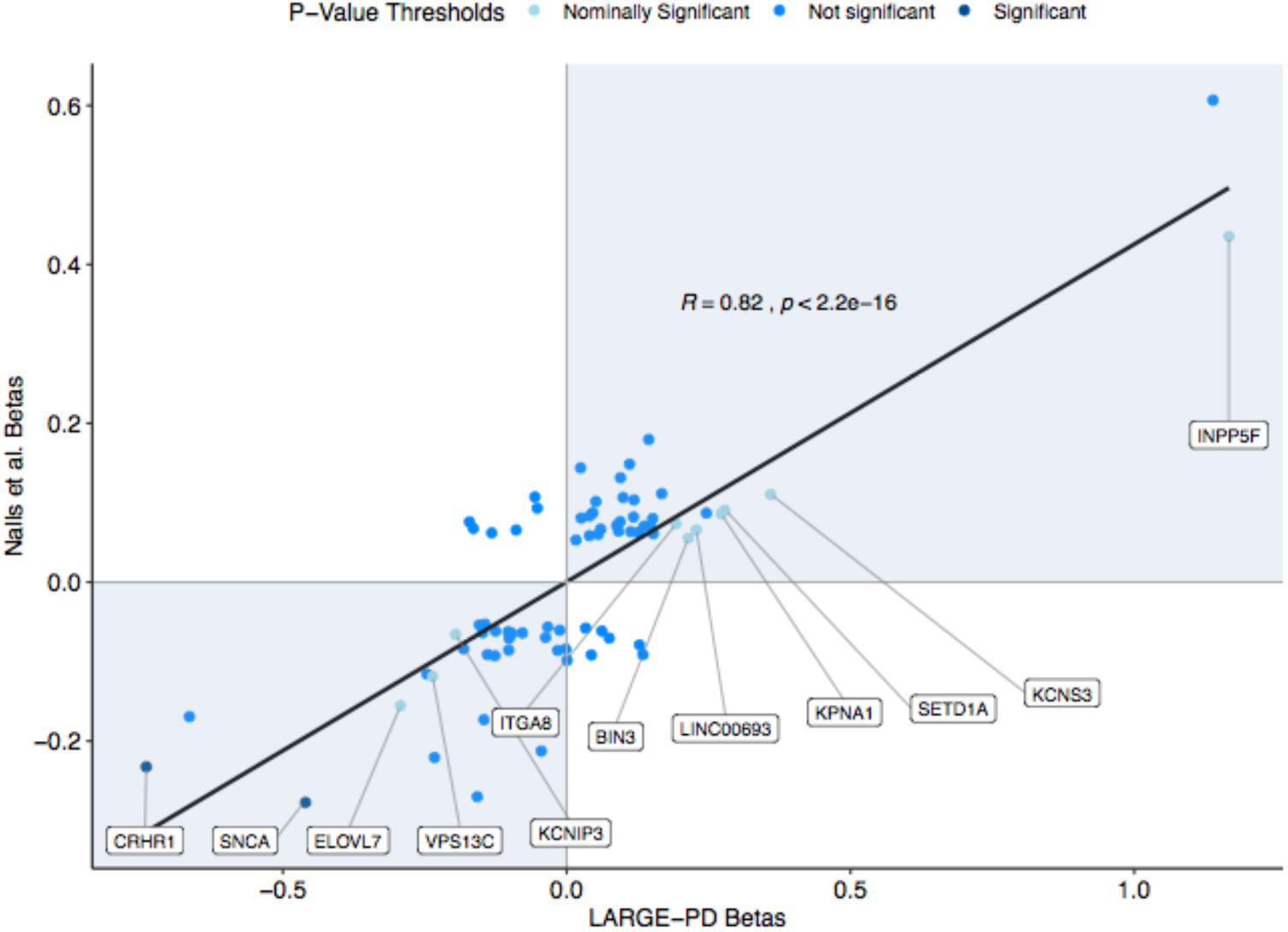
Replication of GWAS significant results from Nalls et al. 2019^1^. On the scatterplot of beta coefficients, the x-axis corresponds to betas obtained from LARGE-PD and the y-axis corresponds to beta coefficients from Nalls et al. 2019^1^ for 76 of the 90 GWAS significant variants. In LARGE-PD, we successfully imputed 84 of the 90 variants; this figure excludes three variants with a MAC less than 10 and five strand ambiguous (CG/AT) sites that did not pass our filters (see methods). The color scheme represents p-values obtained from LARGE-PD. Significant (p-value < 5.9×10^−4^) and nominally significant (p-value < 0.05) variants are labeled by their respective nearest genes.

In addition to the replication of Nalls et al. 2019, we also performed a look-up of PD risk variants from Nalls et al. 2014, Chang et al. 2017, and Foo et al. 2020 (**see supplementary table 6**).^6,7,9^ 36 out of the 41 variants we looked up were consistent in their direction of effect, including both variants from Foo et al. Worth noting, Foo et al. 2020 is a study of individuals with East Asian ancestry, while the other two studies only utilize European-ancestry subjects. We also looked up the three independent PD risk variants in *SNCA* that were identified by Pihlstrøm et al.^44^ One, rs356182, was already included in our replication study. The other two, rs2870004 and rs763443, were not genome-wide significant (p=0.5 and p=0.0015) but were consistent in effect size direction. Neither were in LD with rs356182 in LARGE-PD (R^2^ 0.08 and 0.01, respectively).

### Admixture Mapping

Admixture mapping can be employed if a phenotype shows evidence of differential risk by ancestral background or if we observe allele frequency differences across ancestral populations. For PD, we do see global patterns of PD incidence and prevalence suggestive of differential PD risk.^2^ In addition, Medicaid data in the United States also indicates potential differences in PD risk by ancestral background.^14^ To explore this in LARGE-PD, we tested the ancestry proportions estimated using ADMIXTURE via logistic regression, adjusting for age, sex, and recruitment site. We found that African ancestry was significantly associated with lower PD risk (p-value < 0.05, **see supplementary figure 7**). Given this result, we next performed admixture mapping to test local ancestry blocks for associations with PD risk. To do so, we employed a joint test using the GENESIS package and local ancestry inferred via RFMix and assuming three-way admixture (**see methods**). This was followed by a single-ancestry analysis to determine the ancestry driving each signal. In the joint test, a locus on chromosome 14 was significantly associated with PD risk at a p-value threshold of 5×10^−5^ (**see Figure 5A; Table 2)**. Three other loci, at chromosomes 6, 17, and 21, were approaching the p-value threshold in the joint test. In the single-ancestry tests, the chromosome 6 locus was significant in the African-ancestry model and the chromosome 14 locus was significant in the Native American model (**supplementary figure 8**). The suggestive peaks at chromosome 17 and 21 were also driven by Native American ancestry.

**Table 2:** Admixture mapping results.

| CHR | PEAK | P | P_ADJ | AFR_PVAL | ADJ_AFR_PVAL | NAM_PVAL | ADJ_NAM_PVAL |
| --- | --- | --- | --- | --- | --- | --- | --- |
| 6 | 166465311-166607482 | 1.11E-04 | 0.111 | 2.02E-05 | 0.020 | 0.110 | 1.000 |
| 14 | 24713480-25147976 | 3.57E-05 | 0.036 | 0.012 | 1.000 | 1.91E-05 | 0.019 |
| 17 | 33970400-34509873 | 3.22E-04 | 0.322 | 0.346 | 1.000 | 9.19E-05 | 0.092 |
| 21 | 44767470-46068473 | 2.21E-04 | 0.221 | 0.073 | 1.000 | 6.92E-05 | 0.069 |

LARGE-PD Admixture mapping (AM) results. CHR, the chromosomal local of the AM peak. PEAK, the physical position in build hg19. P and P\_ADJ columns are the p-values from the joint test. The AFR\_PVAL and ADJ\_AFR\_PVAL are the p-values for the African single-ancestry test; the NAM\_PVAL and ADJ\_NAM\_PVAL are
the p-values for the Native American single-ancestry test. All adjusted p-values (ADJ) are corrected for a significance threshold of 5E-05 (see supplementary methods).

**Figure 5:**
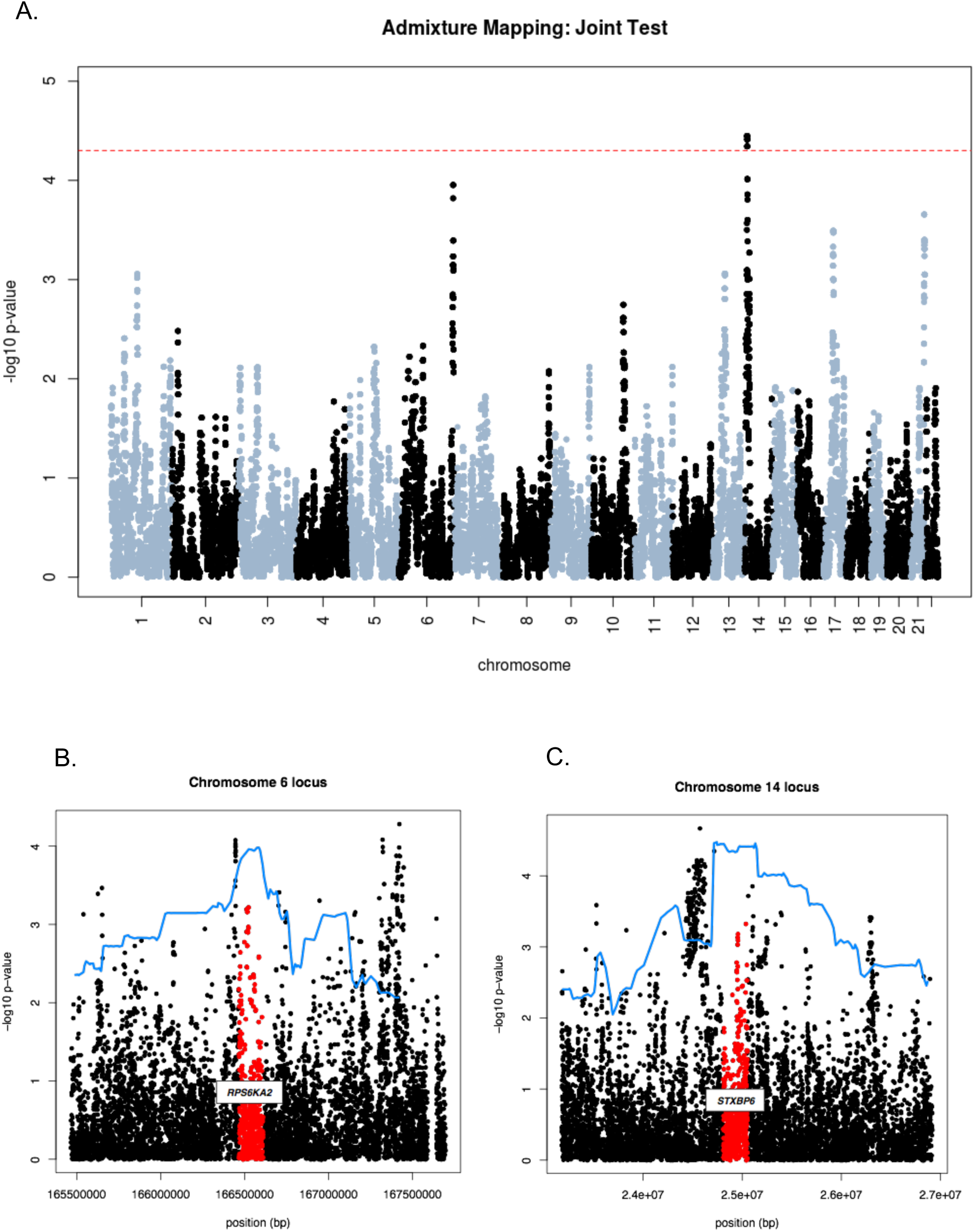
LARGE-PD Admixture mapping results. 5A: Admixture mapping result of a joint test as implemented by the GENESIS package in R. The significance level of 5×10^−5^ is indicated in red. 5B and 5C: The admixture mapping results (blue) are fit using Loess curve and overlaid on the GWAS results in that region. The gene co-localized with the admixture mapping peak is labeled and highlighted in red.

To fine-map the admixture mapping signal, we tested imputed variants co-localized within each peak using a logistic mixed model and evaluated their significance using a regional significance level determined by the number of imputed variants within each peak **(see supplementary table 7)**. For two of the peaks, chromosome 6 and 21, the SNP with the lowest p-value is intronic; for the other two peaks, the SNP with the lowest p-value is intergenic. The chromosome 6 admixture mapping peak contains *RPS6KA2* (**Figure 5B)**; an intronic variant, rs75880521, achieved the lowest p-value (6.05×10^−4^), but this was not significant when adjusting for the number of variants in the region (adjusted p-value of 0.35). This variant had a MAF of 0.22 in Africans in 1000 Genomes but is virtually absent in populations without African ancestry. The chromosome 14 locus encompasses *STXBP6* (**Figure 5C)** and rs79647551 achieved the lowest p-value (4.5 ×10^−5^). This variant is intergenic and had a frequency of 0.31 in Admixed Latin American populations but was considerably less frequent in other populations. The chromosome 17 locus does not contain a gene; within this locus, rs4795926 had the lowest p-value (1.7×10^−3^). This variant was most frequent in Admixed Latin American populations. The chromosome 21 locus encompasses *ITGB2* where rs183517, an intronic variant, had a p-value of 1.6×10^−4^. In 1000 Genomes, this variant had the lowest frequency in Admixed American populations.

## Discussion

As has been well-documented by Sirugo et al. 2019, the majority of GWAS subjects represented in the GWAS catalog are of European ancestry.^11^ PD GWAS efforts feature a similar disparity; with the exception of three studies in East Asia^8,9,60^, the last three large-scale studies have exclusively included individuals of European descent.^1,6,7^ This risks missing population-specific variation that could impact PD risk and possibly lead to predictive disparities through the use of polygenic risk scores.^11,61^ LARGE-PD, the first Latino PD GWAS cohort, is indicative of a shift towards more inclusive PD genomic research.

In LARGE-PD, we estimated the additive heritability of PD to be 0.38 (SE 0.068) with a prevalence of 0.5%. The heritability estimate, though higher than that of European cohorts^62^, was concordant with a study of Kaiser Permanente members where more familial aggregation of PD was observed in Hispanics/Latinos that in other population classifications.^63^ Nevertheless, further refinement with a larger sample size is necessary to improve the accuracy of PD heritability estimation in South American populations.

The suggestive chromosome 3 locus identified by our GWAS was driven by Peruvians of primarily Amerindian ancestry (**Figure 1)**. The nearest gene to this locus, *NRROS* (Negative Regulator Of Reactive Oxygen Species; also called *LRRC33*), is biologically plausible as a potential PD risk gene. *NRROS* knockout mice display neurological abnormalities including motor deficits^53^ and a neurodegenerative phenotype has recently been identified in patients who are homozygous for loss-of-function *NRROS* variants.^64^ In addition, *NRROS* appears to be critical for microglial development.^53,65^ However, the chromosome 3 locus did not replicate in the 23andMe cohort nor was it nominally significant, so it is possible that this was simply a false positive. However, the mean ancestral proportions of 23andMe differs from that of LARGE-PD, with a mean Native American ancestry of 19% (16.5% in cases) in 23andMe compared to a mean Amerindian ancestry of 46.9% in LARGE-PD and 78.3% in the Peru-Lima subset (**see supplementary table 1)**. To determine if this locus was truly a false positive, an additional replication in a cohort with greater Amerindian ancestry might be necessary.

The *SNCA* locus in chromosome 4 achieved genome-wide significance in both LARGE-PD and the 23andMe replication cohort (**Table 1**). In large-scale PD GWAS and meta-analyses, the strongest associations were consistently within the *SNCA* locus, though such studies have been limited to populations of European and East Asian ancestry.^44,55–58^ In LARGE-PD, 28 variants in *SNCA* achieved genome-wide significance, with 20 replicating in 23andMe. This includes rs356182, the lead variant at the *SNCA* locus in the European-ancestry studies. Twenty-five of the variants were represented in data available in the PDGene portal^66^; all 25 were genome-wide significant in PDGene. The 28 variants formed three blocks of tight LD (**Figure 3)** in LARGE-PD. In our conditional analysis, there appeared to be two independent signals, with one being rs356182, when we corrected for the 28 variants. However, if we utilized the more stringent regional correction employed by Pihlstrøm et al.^44^, then we see minimal evidence of a signal independent from rs356182, though our analysis likely lacked power. Worth noting is the higher LD between *SNCA* variants in the Peruvian subjects who made up over half of LARGE-PD subjects (**supplementary figure 4**). Fourteen of the 28 significant variants were tightly correlated with rs356182 (R^2^ > 0.8), the lead *SNCA* SNP in European PD studies, and all tested variants were at least moderately correlated with rs356182 in Peruvian subjects (R^2^ > 0.41). This suggests that the signal we observed in the *SNCA* locus was indeed being driven by rs356182. In addition, rs356182 was the only variant that was genome-wide significant in both LARGE-PD and the 23andMe replication cohort. A regional stepwise conditional analysis in a large diverse dataset is necessary to determine the number of independent PD risk variants in *SNCA*. Nevertheless, it is clear that *SNCA*, and rs356182 in particular, plays a significant role in PD etiology in Latinos.

In our replication of the independent GWAS-significant variants identified by Nalls et al.^1^, we found that 82% of the tested variants were concordant in their effect size direction in LARGE-PD. Two of the variants, rs356182 (*SNCA*) and rs117615688 (nearest gene *CRHR1* in the *MAPT* locus) replicated, with rs356182 achieving genome-wide significance. In our look-up of PD risk variants with a minimum MAF of 1% across three studies,^6,7,9^ 34 out of the 39 variants identified in European studies were concordant in their effect size direction; both of the variants identified in an East Asian cohort were also concordant. Despite challenges estimating beta coefficients due to sample size, we found evidence that there is a substantial overlap in the genetic architecture of PD between Latinos and Europeans.

In our exploration of the relationship between ancestry and PD risk, we found evidence that African ancestry was protective against PD risk (**supplementary figure 7**) and there was a statistically-significant locus on chromosome 6 in the African-ancestry admixture mapping model (**supplementary figure 8C**). Fine-mapping the chromosome 6 locus found rs58837225, an intronic variant in *RPS6KA2* (Ribosomal Protein S6 Kinase A2) that was common in individuals of African ancestry but rare in other populations. A variant in *RPS6KA2* was recently shown to be in an three-way epistatic relationship with a variant in *SNCA* and a variant in *RPTOR* in an age at PD onset study.^67^ However, this admixture peak did not achieve significance in the joint test nor was the fine-mapped variant regionally significant (**see methods**). The mean proportion of African ancestry in LARGE-PD was under 0.06, meaning that we were underpowered to detect African-specific variation. A second locus on chromosome 14 achieved significance in the joint test and in the Native American-ancestry model (**Figure 5A, 5C, supplementary figure 8B**). This locus contains the gene *STXBP6* (Syntaxin Binding Protein 6); though it has primarily been implicated in lung cancer^68^, this gene is also highly expressed in the brain.^69^ Though our admixture mapping results likely provide information for hypothesis generation, replication of our results is necessary, ideally in a cohort enriched in African ancestry for the chromosome 6 result and a cohort enriched in Native American ancestry for our chromosome 14 result.

LARGE-PD is a significant step forward towards increasing the diversity in PD GWAS efforts, though a comprehensive understanding of population-specific PD genetic architecture is still lacking outside of individuals of European and East Asian origin. Larger sample sizes are always needed, but just as necessary is the inclusion of Hispanic/Latino PD subjects from diverse ancestral backgrounds, such as those with significant Native American or African ancestries. PD is a global disease, and it is crucial that genetic studies reflect a wide diversity of individuals.

## Data Availability

The full LARGE-PD GWAS summary statistics are available upon request. Summary statistics for the 23andMe replication dataset are given in the manuscript.

## Acknowledgments

We thank all of the individuals who participated in LARGE-PD. We also want to thank all the support staff at the different Latin American sites for their efforts and support building this incredible resource. Finally, we would like to thank the research participants and employees of 23andMe for making this work possible. Members of the 23andMe Research Team: Michelle Agee, Stella Aslibekyan, Adam Auton, Robert K. Bell, Katarzyna Bryc, Sarah K. Clark, Sarah L. Elson, Kipper Fletez-Brant, Pierre Fontanillas, Nicholas A. Furlotte, Pooja M. Gandhi, Barry Hicks, David A. Hinds, Karen E. Huber, Ethan M. Jewett, Yunxuan Jiang, Aaron Kleinman, Keng-Han Lin, Nadia K. Litterman, Marie K. Luff, Jennifer C. McCreight, Matthew H. McIntyre, Kimberly F. McManus, Joanna L. Mountain, Sahar V. Mozaffari, Priyanka Nandakumar, Elizabeth S. Noblin, Carrie A.M. Northover, Jared O’Connell, Aaron A. Petrakovitz, Steven J. Pitts, G. David Poznik, J. Fah Sathirapongsasuti, Janie F. Shelton, Suyash Shringarpure, Chao Tian, Joyce Y. Tung, Robert J. Tunney, Vladimir Vacic, Xin Wang.

## Appendix

Members of the Latin American Research Consortium on the Genetics of PD (LARGE-PD): Argentina: Federico Micheli, Emilia Gatto.

Brazil: Vitor Tumas, Vanderci Borges, Henrique B. Ferraz, Carlos R.M. Rieder, Artur Schumacher-Schuh, Bruno L. Santos-Lobato.

Chile:Pedro Chaná.

Colombia: Carlos Velez-Pardo, Marlene Jimenez-Del-Rio, Francisco Lopera, Gonzalo Arboleda, Humberto Arboleda, Jorge Luis Orozco, Sonia Moreno, William Fernandez, Carlos E. Arboleda-Bustos.

Costa Rica:Jaime Fornaguera, Alvaro Hernández Guillén, Gabriel Torrealba Acosta.

Ecuador: Jorge Chang-Castello, Brennie Andreé Muñoz.

Honduras: Alex Medina, Anabelle Ferrera.

Mexico: Daniel Martinez-Ramirez, Mayela Rodriguez.

Peru: Mario Cornejo-Olivas, Pilar Mazzetti, Hugo Sarapura, Andrea Rivera, Luis Torres, Carlos Cosentino, Angel Medina.

Puerto Rico: Angel Viñuela.

Uruguay: Elena Dieguez, Victor Raggio, Andres Lescano, Ignacio Amorín.

