## Supplemental Methods and Data for "Characterizing the genetic architecture of Parkinson’s disease in Latinos"

### Supplementary Methods

*Principal Components Analysis for Genome-wide Association Study*

We performed principal component analysis (PCA) on all LARGE-PD subjects using the PC-AiR[32] and PC-Relate[33] methods that are implemented in the GENESIS package and is available from Bioconductor.[34] We first estimated a kinship matrix using KING-robust[38], which is robust to unknown population structure. Using measures of ancestry divergence calculated from the KING-robust outputs, PC-AiR partitions the samples into a mutually unrelated and ancestry-representative set of subjects, as well as a related set. PC-AiR performs standard principal component (PC) analysis on the set of unrelated individuals and then projects the components of variation for the related set. We then estimated a kinship matrix using PC-Relate with adjustment of the ancestry-representative principal components derived by PC-AiR to appropriately estimate kinship among sampled individuals in the presence of population structure. We then performed a second round of analyses of PC-AiR and PC-Relate using the kinship matrix obtained in the previous step in order to generate the final principal components.

*Determination of 95% Credible Set*

We determined the 95% credible set using PAINTOR 3.0[43]. PAINTOR provides the probability that a variant is causal given its membership in an annotation set. We selected annotations using neuronal and brain as keywords. We tested each annotation separately; we then tested the log Bayes factor of each model against the base model without annotations using a likelihood ratio test (LRT). This LRT follows a $\chi 2$ distribution with a single degree of freedom. We retained annotations with a p-value less than 0.05 for the final model. After running the final model, variants were ordered by their posterior probabilities. We determined the credible set by selecting the variants where the summation of their ordered posterior probabilities is equal to or less than 95% of the total posterior probability.

*23andMe Replication Quality Control*

23andMe association results were extensively QCed following established 23andMe protocol.

Genotyped variants were flagged if they had a Hardy-Weinberg $P < 10^{-20}$, a call rate less than 90%, an association with genotype date with $P < 10^{-50}$, age and sex effects ($r^2 > 0.1$), or failed a test for parent-offspring transmissions. Imputed variants were flagged if they had an imputed $r^2 < 0.3$ or had evidence of a platform batch effect as assessed by an ANOVA of the SNP dosages against the genotype platform ($P < 10^{-50}$). For all variants, variants were flagged if they were only available in 20% or less of the total dataset or the logistic regression model did not converge.

*Principal Component and Genetic Relatedness Matrices for Admixture Mapping*

We obtained principal components and the genetic relatedness matrix (GRM) using the PC-AiR[32] and PC-Relate[33] methods. We included reference samples from the HapMap project phase III[46] and from the HGDP project (http://www.hagsc.org/hgdp/files.html) in the PC-AiR analysis for improved inference of population structure. We estimated the GRM using PC-Relate with adjustment of the ancestry representative principal components derived by PC-AiR. We then performed a second round of analyses of PC-AiR and PC-Relate using the relatedness matrix obtained in the previous step in order to get final the principal components and an ancestry adjusted GRM to be used in the admixture analyses to adjust for population stratification and account for relatedness.

*Admixture Mapping Model Description*

The full admixture mapping logistic mixed model can be described by:

$$\boldsymbol{logit}(\boldsymbol{\pi}) = X\alpha + \boldsymbol{A_j}\boldsymbol{\beta_j} + \boldsymbol{g} ,$$

where $\pi = P(y = 1 | \boldsymbol{X}, \boldsymbol{A_j}, \boldsymbol{g})$ represents the $N \times 1$ column vector of probabilities of being affected for the $N$individuals conditional to the covariates, local ancestry calls and random effects, $\boldsymbol{X}$ is the matrix of covariates and $\alpha$ is the vector of fixed covariate effects including an intercept. We assume that $\boldsymbol{g} \sim N(0, \sigma_a^2 \Phi)$ is a vector of random effects for the $N$ subjects, where $\sigma_a^2$ is the additive genetic variance and $\Phi$ is the genetic relatedness matrix. Letting the third ancestral population be the reference population, $\boldsymbol{A_j}$ represents a $N \times (K-1)$ matrix of the local ancestry dosages at the locus $j$ for the $K-1$parental populations, with the corresponding effect size vector $\beta_{\boldsymbol{j}}$ of length $K-1$. The null hypothesis that $\beta_{\boldsymbol{j}} = \mathbf{0}$ was assessed via multivariate score test.

*Determination of Admixture Mapping Significance Level*

Based on previous studies, a nominal p-value of $5 \times 10^{-5}$ controls the type I error at level of 0.05.[52] Recent admixture, such as observed in Latinos, creates long-range linkage disequilibrium and thus the significance threshold does not need to be as stringent as that used for the association mapping. We also estimated the significance level empirically by fitting an autoregressive model to the admixture mapping p-values, summing the results across each chromosome.[70,71] From the empirical autoregression model, we obtained a significance level of $7.7 \times 10^{-5}$. We elected to utilize the more conservative p-value threshold of $5 \times 10^{-5}$.

### Supplementary Table 1: Cohort Description

| SITE | N | N CASES | MEAN AGE | SD AGE | SEX RATIO | MEAN AGE ONSET | SD AGE ONSET |
|---|---|---|---|---|---|---|---|
| **Brazil: Porto Alegre** | 13 | 3 | 64.46 | 9.12 | 0.15 | 64.33 | 10.02 |
| **Brazil: Ribeirao Preto** | 195 | 126 | 57.15 | 15.12 | 0.49 | 51.13 | 14.13 |
| **Brazil: Sao Paulo** | 19 | 19 | 59.37 | 9.04 | 0.74 | 50.21 | 9.93 |
| **Chile** | 13 | 13 | 63.67 | 9.59 | 0.75 | 56.83 | 10.06 |
| **Colombia: Bogota** | 23 | 23 | 49.09 | 10.18 | 0.39 | 40.70 | 10.22 |
| **Colombia: Medellin** | 328 | 134 | 60.34 | 12.44 | 0.44 | 51.83 | 15.70 |
| **Peru: Lima** | 670 | 437 | 60.15 | 13.57 | 0.46 | 56.77 | 13.68 |
| **Peru: Puno** | 45 | 0 | 43.00 | 19.80 | 0.24 | NA | NA |
| **Uruguay** | 191 | 52 | 61.13 | 12.62 | 0.35 | 50.77 | 14.51 |
| **TOTAL** | 1497 | 807 | 59.30 | 13.90 | 0.44 | 54.09 | 14.35 |
| **TOTAL: CASES** | 807 | NA | 61.70 | 12.81 | 0.53 | 54.09 | 14.35 |
| **TOTAL: CONTROLS** | 690 | NA | 56.48 | 14.59 | 0.33 | NA | NA |

Description of LARGE-PD cohort. SITE, recruitment site. N, sample size from each site. N CASES, number of cases from each site. MEAN AGE, mean age at data collection. SD AGE, standard deviation of age at data collection. SEX RATIO, the proportion of the cohort that is male. MEAN AGE ONSET, the mean age at disease diagnosis. SD AGE ONSET, the standard deviation of age at onset.

### Supplementary Table 2: Cohort Ancestry

| SITE | MEAN AFR | IQR AFR | MEAN EUR | IQR EUR | MEAN NAT_AMR | IQR NAT_AMR | MEAN OTHER | IQR OTHER |
|---|---|---|---|---|---|---|---|---|
| **Brazil: Porto Alegre** | 0.080 | 0.047 | 0.821 | 0.126 | 0.095 | 0.063 | 0.004 | 0.002 |
| **Brazil: Ribeirao Preto** | 0.092 | 0.109 | 0.825 | 0.208 | 0.063 | 0.093 | 0.020 | 0.007 |
| **Brazil: Sao Paulo** | 0.143 | 0.169 | 0.757 | 0.198 | 0.096 | 0.104 | 0.004 | 0.004 |
| **Chile** | 0.016 | 0.006 | 0.614 | 0.116 | 0.340 | 0.150 | 0.030 | 0.012 |
| **Colombia: Bogota** | 0.025 | 0.012 | 0.596 | 0.068 | 0.374 | 0.080 | 0.005 | 0.010 |
| **Colombia: Medellin** | 0.095 | 0.102 | 0.645 | 0.190 | 0.260 | 0.117 | 0.001 | 0.000 |
| **Peru: Lima** | 0.021 | 0.016 | 0.188 | 0.217 | 0.783 | 0.256 | 0.009 | 0.003 |
| **Peru: Puno** | 1.000E-05 | 0.000 | 6.70E-03 | 0.00 | 0.993 | 0.000 | 2.00E-05 | 0.000 |
| **Uruguay** | 0.051 | 0.034 | 0.825 | 0.176 | 0.120 | 0.137 | 0.004 | 0.002 |
| **All Sites** | 0.052 | 0.060 | 0.472 | 0.629 | 0.469 | 0.694 | 0.008 | 0.002 |

Ancestry composition of LARGE-PD as inferred using ADMIXTURE, a K of 5, and 1000 Genomes Project subjects as references. SITE, recruitment site. MEAN AFR and IQR AFR, the mean and interquartile range of African ancestry proportions. MEAN EUR and IQR EUR, the mean and interquartile range of European ancestry proportions. MEAN NAT_AMR and IQR NAT_AMR, the mean and interquartile range of Native American ancestry proportions. MEAN OTHER and IQR OTHER, the mean and interquartile range of the combination of the inferred East Asian and South Asian components.

### Supplementary Table 3: Conditional analysis results

| SNP | PVAL | P_rs356182 | P_rs356182 | P_rs356182 | P_rs6830166 | P_rs6830166 | P_rs6830166 |
|---|---|---|---|---|---|---|---|

| | | | ADJ | ADJ_REGION | | ADJUSTED | ADJ_REGION |
|---|---|---|---|---|---|---|---|
| rs356183 | 1.120E-08 | 0.044 | 1.000 | 1.000 | 0.396 | 1.000 | 1.000 |
| rs356182 | 4.900E-08 | NA | 1.000 | 1.000 | NA | NA | NA |
| rs356181 | 4.940E-08 | 0.105 | 1.000 | 1.000 | 0.649 | 1.000 | 1.000 |
| rs356211 | 5.230E-08 | 0.068 | 1.000 | 1.000 | 0.921 | 1.000 | 1.000 |
| rs356219 | 4.420E-08 | 0.132 | 1.000 | 1.000 | 0.901 | 1.000 | 1.000 |
| rs356220 | 3.000E-08 | 0.097 | 1.000 | 1.000 | 0.978 | 1.000 | 1.000 |
| rs356221 | 7.660E-09 | 0.019 | 0.524 | 1.000 | 0.544 | 1.000 | 1.000 |
| rs356223 | 5.860E-09 | 0.015 | 0.431 | 1.000 | 0.493 | 1.000 | 1.000 |
| rs356225 | 5.390E-09 | 0.014 | 0.402 | 1.000 | 0.475 | 1.000 | 1.000 |
| rs356165 | 1.520E-08 | 0.058 | 1.000 | 1.000 | 0.798 | 1.000 | 1.000 |
| rs356204 | 6.990E-09 | 0.017 | 0.480 | 1.000 | 0.519 | 1.000 | 1.000 |
| rs356203 | 2.480E-08 | 0.083 | 1.000 | 1.000 | 0.917 | 1.000 | 1.000 |
| rs356200 | 6.990E-09 | 0.017 | 0.480 | 1.000 | 0.519 | 1.000 | 1.000 |
| rs189596 | 5.820E-09 | 0.015 | 0.420 | 1.000 | 0.487 | 1.000 | 1.000 |
| rs356168 | 5.820E-09 | 0.015 | 0.420 | 1.000 | 0.487 | 1.000 | 1.000 |
| rs2736990 | 5.820E-09 | 0.015 | 0.420 | 1.000 | 0.487 | 1.000 | 1.000 |
| rs356198 | 6.920E-08 | 0.002 | 0.062 | 0.496 | 0.026 | 0.728 | 1.000 |
| rs356197 | 6.920E-08 | 0.002 | 0.062 | 0.496 | 0.026 | 0.728 | 1.000 |
| rs356196 | 6.920E-08 | 0.002 | 0.062 | 0.496 | 0.026 | 0.728 | 1.000 |
| rs356191 | 2.630E-08 | 1.20E-03 | 0.035 | 0.278 | 0.019 | 0.526 | 1.000 |
| rs356162 | 3.250E-08 | 1.40E-03 | 0.039 | 0.314 | 0.021 | 0.591 | 1.000 |
| rs184810 | 3.250E-08 | 1.40E-03 | 0.039 | 0.314 | 0.021 | 0.591 | 1.000 |
| rs3775434 | 6.920E-09 | 5.00E-04 | 0.013 | 0.103 | 0.690 | 1.000 | 1.000 |
| rs3822089 | 9.470E-09 | 6.00E-04 | 0.017 | 0.132 | 0.915 | 1.000 | 1.000 |
| rs3822090 | 9.470E-09 | 6.00E-04 | 0.017 | 0.132 | 0.915 | 1.000 | 1.000 |
| rs3775439 | 2.200E-08 | 1.10E-03 | 0.030 | 0.241 | 0.448 | 1.000 | 1.000 |
| rs2737029 | 1.920E-08 | 0.032 | 0.900 | 1.000 | 0.984 | 1.000 | 1.000 |
| rs6830166 | 6.530E-09 | 4.00E-04 | 0.012 | 0.096 | NA | NA | NA |

SNP, the rs ID of the variant. PVAL, the p-value prior to conditioning. P_rs356182, P_rs356182 ADJ, and P_rs356182 ADJ_REGION: p-values when conditioning on rs356182. P_rs6830166, P_rs6830166 ADJUSTED, and P_rs6830166 ADJ_REGION: p-values when conditioning on rs356182 and rs6830166. ADJ p-values correct for the number of GWAS-significant variants (28) and ADJ_REGION corrects for 220 tests (see methods).

### Supplementary Table 4: 23andMe Replication of LARGE-PD results

| SNP | CHROM | POS | ALLELES | GENE.CONTEXT | PVALUE | BETA | SE |
|---|---|---|---|---|---|---|---|
| rs271327 | chr1 | 30115843 | A/G | PTPRU---[] | 0.932 | 0.006 | 0.066 |
| rs9701252 | chr1 | 211041590 | C/T | [KCNH1] | 0.895 | -0.008 | 0.058 |
| rs7517868 | chr1 | 224251666 | A/G | AC138393.1--[]--FBXO28 | 0.023 | 0.097 | 0.043 |
| rs708736 | chr1 | 226709895 | A/G | PARP1---[]--STUM | 0.157 | 0.069 | 0.049 |
| rs2759345 | chr1 | 231948939 | C/T | [TSNAX-DISC1,DISC1] | 0.059 | -0.081 | 0.043 |
| rs17700588 | chr2 | 18717735 | C/G | KCNS3---[]--RDH14 | 0.286 | -0.067 | 0.063 |
| rs13418209 | chr2 | 164727379 | G/T | FIGN---[]---GRB14 | 0.767 | 0.014 | 0.046 |
| rs79278178 | chr2 | 182609741 | G/T | NEUROD1--[]---SSFA2 | 0.104 | -0.258 | 0.165 |
| rs734184 | chr3 | 59554896 | A/G | C3orf67---[]---FHIT | 0.774 | -0.012 | 0.042 |
| rs1711557 | chr3 | 99425669 | A/G | [COL8A1] | 0.327 | 0.049 | 0.050 |
| rs62410627 | chr3 | 196323379 | A/G | FBXO45-[]--NRROS | 0.894 | 0.012 | 0.088 |

| | | | | | | | |
|---|---|---|---|---|---|---|---|
| rs75566098 | chr3 | 196327220 | C/T | FBXO45--[]--NRROS | 0.855 | -0.015 | 0.084 |
| rs112491204 | chr3 | 196327802 | A/G | FBXO45--[]--NRROS | 0.851 | -0.016 | 0.084 |
| rs4916439 | chr3 | 196327848 | A/G | FBXO45--[]--NRROS | 0.850 | -0.016 | 0.085 |
| rs4916525 | chr3 | 196327876 | C/T | FBXO45--[]--NRROS | 0.824 | -0.019 | 0.085 |
| rs77286998 | chr3 | 196328485 | A/C | FBXO45--[]--NRROS | 0.874 | -0.013 | 0.084 |
| rs144236972 | chr3 | 196328938 | C/T | FBXO45--[]--NRROS | 0.861 | -0.015 | 0.084 |
| rs113962850 | chr3 | 196329372 | A/G | FBXO45--[]--NRROS | 0.808 | -0.020 | 0.084 |
| rs111874183 | chr3 | 196330508 | C/T | FBXO45--[]--NRROS | 0.842 | -0.017 | 0.084 |
| rs74394036 | chr3 | 196331602 | C/T | FBXO45--[]--NRROS | 0.854 | 0.015 | 0.084 |
| rs113315229 | chr3 | 196331728 | C/T | FBXO45--[]--NRROS | 0.845 | -0.016 | 0.084 |
| rs74507992 | chr3 | 196332063 | A/G | FBXO45--[]--NRROS | 0.853 | -0.016 | 0.084 |
| rs77270731 | chr3 | 196334530 | A/G | FBXO45--[]--NRROS | 0.840 | -0.017 | 0.084 |
| rs78820950 | chr3 | 196357126 | A/T | FBXO45--[]-NRROS | 0.656 | -0.051 | 0.116 |
| rs78345302 | chr3 | 196359181 | C/G | FBXO45--[]-NRROS | 0.875 | -0.017 | 0.106 |
| rs112565872 | chr3 | 196359973 | C/T | FBXO45--[]-NRROS | 0.288 | -0.096 | 0.091 |
| rs112243434 | chr3 | 196361847 | C/T | FBXO45--[]-NRROS | 0.251 | -0.102 | 0.090 |
| rs113727629 | chr3 | 196361884 | G/T | FBXO45--[]-NRROS | 0.251 | -0.102 | 0.090 |
| rs78832270 | chr3 | 196372937 | A/G | [NRROS] | 0.498 | 0.099 | 0.148 |
| rs60523751 | chr4 | 3726293 | A/G | LRPAP1---[]--ADRA2C | 0.905 | 0.019 | 0.156 |
| rs73258550 | chr4 | 22075244 | A/T | KCNIP4---[]---ADGRA3 | 0.166 | 0.130 | 0.096 |
| rs147853385 | chr4 | 90516997 | G/T | GPRIN3---[]---SNCA | 0.127 | -0.161 | 0.108 |
| rs78448001 | chr4 | 90520012 | A/G | GPRIN3---[]---SNCA | 0.126 | -0.144 | 0.096 |
| rs10012402 | chr4 | 90546806 | C/T | GPRIN3---[]--SNCA | 0.225 | 0.094 | 0.078 |
| rs1430961 | chr4 | 90552920 | A/G | GPRIN3---[]--SNCA | 0.008 | 0.162 | 0.060 |
| rs12498987 | chr4 | 90554975 | A/T | GPRIN3---[]--SNCA | 0.166 | 0.091 | 0.066 |
| rs28419161 | chr4 | 90566935 | C/T | GPRIN3---[]--SNCA | 0.086 | -0.141 | 0.083 |
| rs7674594 | chr4 | 90579508 | A/G | GPRIN3---[]--SNCA | 0.090 | 0.098 | 0.058 |
| rs356182 | chr4 | 90626111 | A/G | GPRIN3---[]--SNCA | 4.55E-08 | 0.234 | 0.043 |
| rs356211 | chr4 | 90636418 | C/T | GPRIN3---[]-SNCA | 1.04E-06 | -0.205 | 0.042 |
| rs356219 | chr4 | 90637601 | A/G | GPRIN3---[]-SNCA | 2.35E-07 | 0.216 | 0.042 |
| rs11931074 | chr4 | 90639515 | G/T | GPRIN3---[]-SNCA | 3.50E-06 | 0.240 | 0.051 |
| rs356220 | chr4 | 90641340 | C/T | GPRIN3---[]-SNCA | 1.07E-07 | 0.222 | 0.042 |
| rs356221 | chr4 | 90642464 | A/T | GPRIN3---[]-SNCA | 5.48E-07 | -0.209 | 0.042 |
| rs356223 | chr4 | 90643507 | A/G | GPRIN3---[]-SNCA | 6.82E-07 | -0.207 | 0.042 |
| rs356225 | chr4 | 90643757 | C/G | GPRIN3---[]-SNCA | 4.48E-07 | -0.211 | 0.042 |
| rs8180209 | chr4 | 90644454 | A/G | GPRIN3---[]SNCA | 3.38E-06 | 0.240 | 0.051 |
| rs1045722 | chr4 | 90645671 | A/T | [SNCA] | 2.50E-06 | -0.245 | 0.051 |
| rs3857053 | chr4 | 90645674 | C/T | [SNCA] | 3.37E-06 | 0.241 | 0.051 |
| rs356165 | chr4 | 90646886 | A/G | [SNCA] | 8.94E-08 | 0.224 | 0.042 |
| rs356204 | chr4 | 90663542 | C/T | [SNCA] | 4.26E-07 | 0.211 | 0.042 |
| rs356203 | chr4 | 90666041 | C/T | [SNCA] | 8.14E-08 | -0.224 | 0.042 |
| rs356200 | chr4 | 90668614 | C/T | [SNCA] | 4.00E-07 | 0.211 | 0.042 |
| rs189596 | chr4 | 90671336 | A/G | [SNCA] | 5.34E-07 | 0.209 | 0.042 |
| rs356168 | chr4 | 90674431 | A/G | [SNCA] | 5.61E-07 | 0.209 | 0.042 |
| rs3857059 | chr4 | 90675238 | A/G | [SNCA] | 3.04E-06 | 0.242 | 0.051 |
| rs567871246 | chr4 | 90678255 | A/G | [SNCA] | 4.74E-02 | 0.254 | 0.132 |
| rs2736990 | chr4 | 90678541 | A/G | [SNCA] | 5.13E-07 | 0.209 | 0.042 |
| rs356198 | chr4 | 90682504 | C/T | [SNCA] | 1.50E-03 | -0.167 | 0.053 |
| rs356197 | chr4 | 90682750 | A/G | [SNCA] | 1.49E-03 | 0.167 | 0.053 |
| rs10516845 | chr4 | 90684278 | A/G | [SNCA] | 5.26E-08 | 0.299 | 0.054 |
| rs356191 | chr4 | 90688120 | A/G | [SNCA] | 8.68E-04 | 0.169 | 0.051 |

| | | | | | | | |
|---|---|---|---|---|---|---|---|
| rs356188 | chr4 | 90691537 | C/T | [SNCA] | 9.77E-04 | 0.167 | 0.052 |
| rs356162 | chr4 | 90697157 | C/T | [SNCA] | 8.99E-04 | 0.168 | 0.051 |
| rs184810 | chr4 | 90697979 | C/T | [SNCA] | 9.10E-04 | 0.168 | 0.051 |
| rs3775434 | chr4 | 90702781 | A/G | [SNCA] | 9.25E-06 | 0.218 | 0.048 |
| rs3822089 | chr4 | 90704011 | A/G | [SNCA] | 1.14E-05 | -0.216 | 0.048 |
| rs3822090 | chr4 | 90704876 | C/T | [SNCA] | 1.06E-05 | 0.216 | 0.048 |
| rs3775439 | chr4 | 90709741 | A/G | [SNCA] | 1.19E-05 | -0.215 | 0.048 |
| rs2737030 | chr4 | 90710099 | C/T | [SNCA] | 9.93E-04 | 0.166 | 0.051 |
| rs2737029 | chr4 | 90711770 | C/T | [SNCA] | 4.21E-07 | -0.212 | 0.042 |
| rs10014396 | chr4 | 90712629 | C/T | [SNCA] | 7.46E-05 | -0.220 | 0.054 |
| rs2583962 | chr4 | 90713747 | C/T | [SNCA] | 1.00E-03 | -0.166 | 0.051 |
| rs3889917 | chr4 | 90716852 | C/T | [SNCA] | 1.69E-02 | -0.149 | 0.061 |
| rs2737028 | chr4 | 90717016 | A/G | [SNCA] | 9.81E-04 | 0.166 | 0.051 |
| rs2572318 | chr4 | 90718719 | C/T | [SNCA] | 9.83E-04 | 0.166 | 0.051 |
| rs2737025 | chr4 | 90719192 | A/G | [SNCA] | 9.93E-04 | 0.166 | 0.051 |
| rs2583964 | chr4 | 90727218 | A/G | [SNCA] | 7.23E-04 | 0.170 | 0.051 |
| rs2197120 | chr4 | 90729602 | A/G | [SNCA] | 1.27E-03 | 0.162 | 0.051 |
| rs2619368 | chr4 | 90729747 | G/T | [SNCA] | 1.08E-03 | -0.164 | 0.051 |
| rs748849 | chr4 | 90734961 | A/G | [SNCA] | 1.11E-03 | -0.163 | 0.051 |
| rs1442144 | chr4 | 90736113 | C/T | [SNCA] | 1.10E-03 | -0.163 | 0.051 |
| rs2619341 | chr4 | 90741773 | A/G | [SNCA] | 1.09E-03 | 0.164 | 0.051 |
| rs10005233 | chr4 | 90743331 | C/T | [SNCA] | 1.13E-01 | 0.067 | 0.042 |
| rs10008964 | chr4 | 90744721 | A/G | [SNCA] | 1.15E-03 | -0.163 | 0.051 |
| rs6830166 | chr4 | 90744993 | C/T | [SNCA] | 4.88E-05 | 0.199 | 0.048 |
| rs2619347 | chr4 | 90745770 | A/G | [SNCA] | 8.31E-04 | -0.168 | 0.051 |
| rs2619349 | chr4 | 90746646 | G/T | [SNCA] | 7.65E-04 | 0.169 | 0.051 |
| rs1811442 | chr4 | 90747751 | C/T | [SNCA] | 7.16E-04 | 0.170 | 0.051 |
| rs2619355 | chr4 | 90748374 | A/C | [SNCA] | 7.05E-04 | 0.170 | 0.051 |
| rs2583979 | chr4 | 90750588 | A/T | [SNCA] | 9.60E-04 | 0.167 | 0.051 |
| rs2737002 | chr4 | 90753180 | A/G | [SNCA] | 5.35E-04 | 0.176 | 0.052 |
| rs1471484 | chr4 | 90754313 | C/T | [SNCA] | 5.11E-04 | 0.177 | 0.052 |
| rs990085 | chr4 | 90754771 | A/G | [SNCA] | 5.32E-04 | 0.176 | 0.052 |
| rs1372518 | chr4 | 90757294 | A/C | [SNCA] | 6.39E-04 | 0.171 | 0.051 |
| rs1372519 | chr4 | 90757309 | A/G | [SNCA] | 7.41E-04 | 0.169 | 0.051 |
| rs2301134 | chr4 | 90758945 | A/G | [SNCA] | 8.50E-02 | 0.072 | 0.042 |
| rs6822088 | chr4 | 90764310 | C/T | SNCA-[]--MMRN1 | 6.98E-05 | -0.196 | 0.049 |
| rs6532194 | chr4 | 90780902 | C/T | SNCA--[]--MMRN1 | 0.012 | 0.129 | 0.051 |
| rs114568886 | chr4 | 114427290 | A/G | [CAMK2D] | 0.238 | -0.436 | 0.347 |
| rs6847111 | chr4 | 140693857 | C/T | [MAML3] | 0.980 | -0.001 | 0.049 |
| rs6820425 | chr4 | 183226376 | A/C | [TENM3] | 0.753 | 0.018 | 0.058 |
| rs77572607 | chr5 | 76926748 | A/G | [OTP] | 0.067 | 0.161 | 0.090 |
| rs2368554 | chr5 | 97794710 | A/G | []---RGMB | 0.048 | 0.098 | 0.050 |
| rs106451 | chr5 | 104048870 | A/C | [] | 0.131 | -0.085 | 0.056 |
| rs73781866 | chr5 | 109280421 | C/T | MAN2A1--[]---TMEM232 | 0.879 | 0.013 | 0.088 |
| rs79212064 | chr5 | 115342038 | C/G | [LVRN] | 0.183 | 0.095 | 0.071 |
| rs59521270 | chr5 | 115344826 | C/G | [LVRN] | 0.236 | -0.084 | 0.070 |
| rs7732824 | chr5 | 115349682 | A/G | [LVRN] | 0.522 | 0.046 | 0.071 |
| rs443402 | chr5 | 148818868 | A/G | IL17B--[]--CSNK1A1 | 0.550 | 0.045 | 0.075 |
| rs7741438 | chr6 | 5723118 | A/G | [FARS2] | 0.158 | 0.062 | 0.044 |
| rs9784801 | chr6 | 10572987 | A/G | [GCNT2] | 0.039 | -0.380 | 0.195 |
| rs1264344 | chr6 | 30800577 | C/T | IER3--[]--DDR1 | 0.914 | 0.005 | 0.042 |

| | | | | | | | |
|---|---|---|---|---|---|---|---|
| rs3130787 | chr6 | 30809864 | A/G | IER3--[]--DDR1 | 0.620 | -0.021 | 0.042 |
| rs2394450 | chr6 | 30817501 | A/G | IER3---[]--DDR1 | 0.506 | 0.028 | 0.042 |
| rs3130794 | chr6 | 30832810 | C/T | IER3---[]--DDR1 | 0.942 | 0.003 | 0.042 |
| rs3132576 | chr6 | 30833116 | A/G | IER3---[]--DDR1 | 0.928 | 0.004 | 0.042 |
| rs3095354 | chr6 | 30836111 | C/T | IER3---[]--DDR1 | 0.942 | 0.003 | 0.042 |
| rs3132573 | chr6 | 30840688 | A/G | IER3---[]--DDR1 | 0.901 | -0.005 | 0.042 |
| rs3130796 | chr6 | 30840950 | C/T | IER3---[]--DDR1 | 0.938 | -0.003 | 0.042 |
| rs78809511 | chr6 | 32190969 | C/T | [NOTCH4] | 0.149 | -0.247 | 0.177 |
| rs2296340 | chr6 | 33630472 | A/G | [ITPR3] | 0.236 | 0.085 | 0.072 |
| rs11751469 | chr6 | 33804547 | C/T | MLN--[]---GRM4 | 0.142 | -0.064 | 0.044 |
| rs6901216 | chr6 | 33807091 | C/T | MLN--[]---GRM4 | 0.123 | 0.067 | 0.044 |
| rs6914721 | chr6 | 34438564 | A/G | [PACSIN1] | 0.286 | -0.053 | 0.050 |
| rs3862822 | chr6 | 113979801 | A/G | []---MARCKS | 0.165 | -0.106 | 0.077 |
| rs2882346 | chr6 | 115275562 | A/G | HS3ST5---[]---FRK | 0.683 | -0.018 | 0.044 |
| rs58382092 | chr6 | 117730414 | D/I | [ROS1,RP1-179P9.3] | 0.294 | -0.044 | 0.042 |
| rs9403515 | chr6 | 143908013 | A/G | FUCA2--[]--PHACTR2 | 0.851 | 0.008 | 0.045 |
| rs11155309 | chr6 | 143918183 | C/T | FUCA2--[]--PHACTR2 | 0.881 | 0.006 | 0.044 |
| rs7741506 | chr6 | 143951867 | A/C | FUCA2---[]--PHACTR2 | 0.345 | -0.042 | 0.044 |
| rs56352728 | chr7 | 10413783 | A/G | []---NDUFA4 | 0.300 | 0.125 | 0.123 |
| rs116572254 | chr7 | 41700811 | A/G | SUGCT---[]--INHBA | 0.801 | -0.076 | 0.303 |
| rs4510764 | chr7 | 93136812 | A/C | [CALCR] | 0.193 | -0.058 | 0.045 |
| rs143177239 | chr7 | 118862299 | G/T | ANKRD7---[] | 0.433 | -0.196 | 0.257 |
| rs7357254 | chr7 | 135466740 | C/T | FAM180A--[]---MTPN | 0.988 | 0.001 | 0.051 |
| rs77540070 | chr7 | 147280255 | A/G | [CNTNAP2] | 0.613 | -0.043 | 0.085 |
| rs72710842 | chr9 | 30373761 | C/T | [] | 0.493 | -0.082 | 0.122 |
| rs7920886 | chr10 | 24060582 | C/T | OTUD1---[]---KIAA1217 | 0.507 | -0.126 | 0.186 |
| rs1255396 | chr10 | 43039187 | C/T | []--ZNF33B | 0.311 | -0.045 | 0.044 |
| rs4430448 | chr10 | 87636974 | A/G | [GRID1] | 0.166 | -0.187 | 0.132 |
| rs117304246 | chr10 | 105553341 | A/G | [SH3PXD2A] | 0.342 | -0.119 | 0.127 |
| rs79570991 | chr10 | 120248430 | A/G | FAM204A---[]---PRLHR | 0.894 | -0.028 | 0.208 |
| rs10751550 | chr10 | 128109449 | C/T | ADAM12--[]-C10orf90 | 0.334 | -0.064 | 0.066 |
| rs12414718 | chr10 | 134147172 | A/G | [LRRC27] | 0.098 | 0.143 | 0.088 |
| rs11038673 | chr11 | 45846619 | G/T | SLC35C1--[]--CRY2 | 0.267 | -0.048 | 0.043 |
| rs112703641 | chr11 | 70424012 | A/G | [SHANK2] | 0.024 | 0.288 | 0.123 |
| rs117954227 | chr11 | 87028818 | C/G | [TMEM135] | 0.393 | 0.108 | 0.128 |
| rs9315984 | chr13 | 43639187 | C/T | [DNAJC15] | 0.359 | 0.064 | 0.070 |
| rs61956689 | chr13 | 66545953 | A/G | []---PCDH9 | 0.854 | -0.015 | 0.083 |
| rs4885004 | chr13 | 73093913 | A/G | DACH1---[]---MZT1 | 0.194 | -0.076 | 0.059 |
| rs9585041 | chr13 | 100018237 | C/T | [UBAC2] | 0.947 | 0.003 | 0.046 |
| rs7983036 | chr13 | 100043087 | A/G | UBAC2-[]---TM9SF2 | 0.930 | 0.004 | 0.046 |
| rs7142675 | chr14 | 25012115 | C/T | CMA1--[]--CTSG | 0.419 | 0.035 | 0.044 |
| rs1957523 | chr14 | 25044712 | C/T | [CTSG] | 0.476 | 0.032 | 0.044 |
| rs10498408 | chr14 | 49058396 | C/T | MDGA2---[]---RPS29 | 0.232 | -0.094 | 0.079 |
| rs60148623 | chr14 | 74177514 | A/T | DNAL1-[]PNMA1 | 0.796 | -0.028 | 0.109 |
| rs34891196 | chr14 | 74294148 | C/G | ELMSAN1--[]--PTGR2 | 0.839 | 0.009 | 0.042 |
| rs35483531 | chr14 | 100653772 | C/T | DEGS2--[]--YY1 | 0.904 | 0.006 | 0.049 |
| rs16964068 | chr15 | 37078689 | C/G | [C15orf41] | 0.555 | 0.026 | 0.044 |
| rs148346037 | chr15 | 65792495 | C/G | [DPP8] | 0.314 | -0.309 | 0.296 |
| rs73541708 | chr15 | 92891626 | A/G | SLCO3A1---[]--ST8SIA2 | 0.466 | 0.044 | 0.060 |
| rs1724409 | chr17 | 43740565 | G/T | [CRHR1] | 0.001 | 0.166 | 0.051 |
| rs1805081 | chr18 | 21140432 | C/T | [NPC1] | 0.272 | -0.049 | 0.044 |

| | | | | | | | |
|---|---|---|---|---|---|---|---|
| rs11660252 | chr18 | 49727447 | A/G | []---DCC | 0.161 | 0.120 | 0.087 |
| rs4890995 | chr18 | 73493754 | A/C | SMIM21---[]---ZNF516 | 0.646 | 0.054 | 0.118 |
| rs6510816 | chr19 | 4531364 | C/T | [PLIN5,CTB-50L17.14] | 0.325 | -0.047 | 0.048 |
| rs80037173 | chr19 | 39164081 | A/T | [ACTN4] | 0.850 | 0.016 | 0.087 |
| rs6105604 | chr20 | 16430248 | A/C | [KIF16B] | 0.945 | -0.015 | 0.220 |
| rs2258671 | chr20 | 25275230 | A/G | [PYGB] | 0.490 | -0.245 | 0.342 |
| rs9983667 | chr21 | 22186687 | A/C | []---NCAM2 | 0.620 | -0.039 | 0.078 |
| rs5748663 | chr22 | 17299261 | C/T | [XKR3] | 0.594 | 0.054 | 0.100 |

Results from the 23andMe replication of LARGE-PD results (see methods). SNP, the rs ID of the tested variant. CHROM, the chromosomal location. POS, the hg19 coordinates. ALLELES, the alleles of the variant. GENE.CONTEXT, the nearest genes. PVALUE, the p-value. BETA, the beta coefficient of the variant. SE, the standard error of the beta coefficients.

### Supplementary Table 5: Replication of Nalls et al. 2019:

| SNP | NEAREST GENE | AF_NALLS | AF_LARGEPD | BETA_NALLS | BETA_LARGEPD | BETA CONCORD | CG/AT | P | P_ADJ |
|---|---|---|---|---|---|---|---|---|---|
| rs114138760 | PMVK | 0.011 | 0.003 | 0.281 | -0.112 | FALSE | TRUE | 0.875 | 1 |
| rs35749011 | KRTCAP2 | 0.017 | 0.005 | 0.607 | 1.139 | TRUE | FALSE | 0.058 | 1 |
| rs76763715 | GBAP1 | 0.995 | 0.998 | -0.747 | -1.329 | TRUE | FALSE | 0.128 | 1 |
| rs6658353 | FCGR2A | 0.501 | 0.397 | 0.065 | -0.005 | FALSE | TRUE | 0.952 | 1 |
| rs11578699 | VAMP4 | 0.195 | 0.137 | -0.07 | 0.075 | FALSE | FALSE | 0.531 | 1 |
| rs823118 | NUCKS1 | 0.566 | 0.379 | 0.107 | 0.099 | TRUE | FALSE | 0.245 | 1 |
| rs11557080 | RAB29 | 0.139 | 0.152 | 0.132 | 0.095 | TRUE | FALSE | 0.395 | 1 |
| rs4653767 | ITPKB | 0.720 | 0.725 | 0.083 | 0.040 | TRUE | FALSE | 0.660 | 1 |
| rs10797576 | SIPA1L2 | 0.140 | 0.201 | 0.111 | 0.167 | TRUE | FALSE | 0.098 | 1 |
| rs76116224 | KCNS3 | 0.904 | 0.927 | 0.11 | 0.359 | TRUE | TRUE | 0.027 | 1 |
| rs2042477 | KCNIP3 | 0.242 | 0.381 | -0.066 | -0.196 | TRUE | TRUE | 0.029 | 1 |
| rs11683001 | MAP4K4 | 0.337 | 0.260 | 0.071 | 0.167 | TRUE | TRUE | 0.081 | 1 |
| rs57891859 | TMEM163 | 0.719 | 0.530 | 0.081 | 0.025 | TRUE | FALSE | 0.770 | 1 |
| rs1474055 | STK39 | 0.131 | 0.256 | 0.18 | 0.145 | TRUE | FALSE | 0.127 | 1 |
| rs73038319 | SATB1 | 0.959 | 0.991 | -0.169 | -0.665 | TRUE | FALSE | 0.126 | 1 |
| rs6808178 | LINC00693 | 0.379 | 0.186 | 0.066 | 0.228 | TRUE | FALSE | 0.039 | 1 |
| rs12497850 | IP6K2 | 0.648 | 0.782 | 0.064 | 0.113 | TRUE | FALSE | 0.281 | 1 |
| rs55961674 | KPNA1 | 0.172 | 0.119 | 0.086 | 0.272 | TRUE | FALSE | 0.041 | 1 |
| rs11707416 | MED12L | 0.367 | 0.299 | -0.063 | -0.143 | TRUE | TRUE | 0.112 | 1 |
| rs1450522 | SPTSSB | 0.674 | 0.626 | -0.062 | -0.125 | TRUE | FALSE | 0.138 | 1 |
| rs10513789 | MCCC1 | 0.811 | 0.728 | 0.149 | 0.111 | TRUE | FALSE | 0.230 | 1 |
| rs873786 | GAK | 0.099 | 0.052 | -0.173 | -0.146 | TRUE | FALSE | 0.428 | 1 |
| rs34311866 | TMEM175 | 0.807 | 0.899 | -0.213 | -0.045 | TRUE | FALSE | 0.748 | 1 |
| rs4698412 | BST1 | 0.553 | 0.626 | 0.104 | 0.119 | TRUE | FALSE | 0.166 | 1 |
| rs34025766 | LCORL | 0.159 | 0.163 | -0.084 | -0.181 | TRUE | TRUE | 0.113 | 1 |
| rs6825004 | SCARB2 | 0.691 | 0.523 | 0.062 | -0.074 | FALSE | TRUE | 0.376 | 1 |
| rs4101061 | FAM47E | 0.711 | 0.485 | -0.091 | -0.140 | TRUE | FALSE | 0.121 | 1 |
| rs6854006 | FAM47E-STBD1 | 0.363 | 0.218 | -0.091 | 0.134 | FALSE | FALSE | 0.193 | 1 |
| rs356182 | SNCA | 0.628 | 0.562 | -0.277 | -0.460 | TRUE | FALSE | 2.48E-08 | 2.09E-06 |
| rs13117519 | CAMK2D | 0.174 | 0.110 | 0.088 | 0.046 | TRUE | FALSE | 0.722 | 1 |

| | | | | | | | | | |
|---|---|---|---|---|---|---|---|---|---|
| rs62333164 | CLCN3 | 0.326 | 0.173 | -0.064 | -0.078 | TRUE | FALSE | 0.502 | 1 |
| rs1867598 | ELOVL7 | 0.902 | 0.892 | -0.155 | -0.293 | TRUE | FALSE | 0.026 | 1 |
| rs26431 | PAM | 0.703 | 0.799 | 0.062 | -0.132 | FALSE | TRUE | 0.205 | 1 |
| rs11950533 | C5orf24 | 0.102 | 0.177 | -0.092 | 0.043 | FALSE | FALSE | 0.688 | 1 |
| rs9261484 | TRIM40 | 0.245 | 0.404 | -0.064 | -0.148 | TRUE | FALSE | 0.096 | 1 |
| rs12528068 | RIMS1 | 0.284 | 0.166 | 0.066 | -0.089 | FALSE | FALSE | 0.433 | 1 |
| rs997368 | FYN | 0.805 | 0.797 | 0.071 | 0.088 | TRUE | FALSE | 0.409 | 1 |
| rs75859381 | RPS12 | 0.967 | 0.894 | -0.221 | -0.233 | TRUE | FALSE | 0.085 | 1 |
| rs199351 | GPNMB | 0.594 | 0.667 | 0.102 | 0.051 | TRUE | FALSE | 0.570 | 1 |
| rs1293298 | CTSB | 0.744 | 0.863 | 0.093 | -0.052 | FALSE | FALSE | 0.680 | 1 |
| rs620513 | FGF20 | 0.268 | 0.290 | -0.086 | -0.102 | TRUE | FALSE | 0.296 | 1 |
| rs2280104 | BIN3 | 0.360 | 0.242 | 0.056 | 0.214 | TRUE | FALSE | 0.025 | 1 |
| rs2086641 | FAM49B | 0.723 | 0.677 | -0.061 | -0.012 | TRUE | FALSE | 0.888 | 1 |
| rs13294100 | SH3GL2 | 0.342 | 0.542 | -0.086 | -0.016 | TRUE | FALSE | 0.854 | 1 |
| rs10756907 | SH3GL2 | 0.767 | 0.754 | -0.093 | -0.126 | TRUE | FALSE | 0.185 | 1 |
| rs6476434 | UBAP2 | 0.734 | 0.789 | -0.062 | 0.062 | FALSE | FALSE | 0.534 | 1 |
| rs896435 | ITGA8 | 0.689 | 0.679 | 0.074 | 0.193 | TRUE | FALSE | 0.028 | 1 |
| rs10748818 | GBF1 | 0.851 | 0.805 | -0.079 | 0.128 | FALSE | FALSE | 0.215 | 1 |
| rs72840788 | BAG3 | 0.216 | 0.124 | 0.076 | 0.094 | TRUE | FALSE | 0.444 | 1 |
| rs117896735 | INPP5F | 0.017 | 0.007 | 0.435 | 1.167 | TRUE | FALSE | 0.010 | 0.852 |
| rs7938782 | RNF141 | 0.878 | 0.894 | 0.087 | 0.246 | TRUE | FALSE | 0.071 | 1 |
| rs12283611 | DLG2 | 0.415 | 0.321 | -0.065 | -0.096 | TRUE | FALSE | 0.289 | 1 |
| rs3802920 | IGSF9B | 0.205 | 0.152 | 0.107 | -0.056 | FALSE | FALSE | 0.630 | 1 |
| rs76904798 | LRRK2 | 0.144 | 0.199 | 0.144 | 0.024 | TRUE | FALSE | 0.811 | 1 |
| rs34637584 | LRRK2 | 0.002 | 0.002 | 2.429 | 0.678 | TRUE | FALSE | 0.412 | 1 |
| rs7134559 | SCAF11 | 0.404 | 0.276 | -0.054 | -0.154 | TRUE | FALSE | 0.092 | 1 |
| rs11610045 | FBRSL1 | 0.490 | 0.238 | 0.06 | 0.056 | TRUE | FALSE | 0.581 | 1 |
| rs9568188 | CAB39L | 0.740 | 0.677 | 0.062 | 0.127 | TRUE | FALSE | 0.147 | 1 |
| rs4771268 | MBNL2 | 0.230 | 0.147 | 0.068 | -0.164 | FALSE | FALSE | 0.160 | 1 |
| rs12147950 | MIPOL1 | 0.438 | 0.444 | -0.053 | -0.144 | TRUE | FALSE | 0.074 | 1 |
| rs11158026 | GCH1 | 0.325 | 0.425 | -0.084 | -0.002 | TRUE | FALSE | 0.984 | 1 |
| rs3742785 | RPS6KL1 | 0.787 | 0.674 | 0.071 | 0.137 | TRUE | FALSE | 0.130 | 1 |
| rs979812 | GALC | 0.442 | 0.516 | 0.061 | 0.153 | TRUE | FALSE | 0.065 | 1 |
| rs2251086 | VPS13C | 0.142 | 0.139 | -0.119 | -0.236 | TRUE | FALSE | 0.045 | 1 |
| rs2904880 | CD19 | 0.309 | 0.533 | -0.065 | -0.104 | TRUE | TRUE | 0.251 | 1 |
| rs11150601 | SETD1A | 0.644 | 0.561 | 0.091 | 0.279 | TRUE | FALSE | 9.81E-04 | 0.082 |
| rs6500328 | NOD2 | 0.599 | 0.659 | 0.059 | 0.040 | TRUE | FALSE | 0.648 | 1 |
| rs3104783 | CASC16 | 0.434 | 0.666 | 0.067 | 0.060 | TRUE | FALSE | 0.502 | 1 |
| rs10221156 | CHD9 | 0.093 | 0.076 | -0.116 | -0.246 | TRUE | FALSE | 0.109 | 1 |
| rs12600861 | CHRNB1 | 0.648 | 0.679 | -0.057 | -0.033 | TRUE | FALSE | 0.705 | 1 |
| rs12951632 | RETREG3 | 0.735 | 0.797 | 0.064 | 0.091 | TRUE | FALSE | 0.360 | 1 |
| rs2269906 | UBTF | 0.653 | 0.486 | 0.063 | 0.128 | TRUE | FALSE | 0.120 | 1 |
| rs850738 | FAM171A2 | 0.606 | 7.00E-01 | -0.071 | -0.101 | TRUE | FALSE | 0.259 | 1 |
| rs62053943 | CRHR1 | 0.155 | 0.081 | -0.27 | -0.158 | TRUE | FALSE | 0.304 | 1 |
| rs117615688 | CRHR1 | 0.067 | 4.500E-02 | -0.232 | -0.741 | TRUE | FALSE | 2.29E-04 | 0.019 |
| rs11658976 | WNT3 | 0.580 | 0.484 | -0.062 | -0.103 | TRUE | FALSE | 0.225 | 1 |
| rs61169879 | BRIP1 | 0.164 | 0.192 | 0.082 | 0.118 | TRUE | FALSE | 0.262 | 1 |
| rs666463 | DNAH17 | 0.833 | 0.913 | 0.076 | -0.171 | FALSE | TRUE | 0.247 | 1 |
| rs1941685 | ASXL3 | 0.498 | 0.707 | 0.053 | 0.016 | TRUE | FALSE | 0.861 | 1 |
| rs12456492 | RIT2 | 0.682 | 0.652 | -0.098 | 0.001 | FALSE | FALSE | 0.993 | 1 |

| | | | | | | | | | |
|---|---|---|---|---|---|---|---|---|---|
| rs8087969 | MEX3C | 0.550 | 0.654 | -0.058 | 0.033 | FALSE | FALSE | 0.696 | 1 |
| rs55818311 | SPPL2B | 0.694 | 0.529 | -0.07 | -0.037 | TRUE | FALSE | 0.672 | 1 |
| rs77351827 | CRLS1 | 0.128 | 0.066 | 0.08 | 0.152 | TRUE | FALSE | 0.367 | 1 |
| rs2248244 | DYRK1A | 0.283 | 0.254 | 0.071 | 0.147 | TRUE | FALSE | 0.117 | 1 |

Replication of the independent loci described in Nalls et al. 2019 using the LARGE-PD GWAS results. SNP, the rs ID. NEAREST GENE, the nearest gene to the variant. AF_NALLS, the allele frequency given by Nalls et al., AF_LARGEPD, the allele frequency in LARGE-PD, BETA_NALLS, the beta coefficients from Nalls et al., BETA_LARGEPD, the beta coefficients from LARGE-PD, BETA CONCORD, indicates consistent direction of effect, CG/AT, indicates whether the variant in strand ambiguous, P, the p-value the LARGE-PD GWAS, P_ADJ, the p-value corrected for 84 tests.

#### Supplementary Table 6: GWAS-significant variant look-up

| STUDY | SNP | GENE(s) | OR_STUDY | OR_LARGEPD | P_LARGE | CONCORDANCE |
|---|---|---|---|---|---|---|
| Nalls et al. 2014 | rs35749011 | GBA/SYT11 | 1.824 | NA | NA | NA |
| Nalls et al. 2014 | rs823118 | RAB7L1/NUCKS1 | 1.122 | 1.104 | 0.245 | TRUE |
| Nalls et al. 2014 | rs10797576 | SIPA1L2 | 1.131 | 1.182 | 0.098 | TRUE |
| Nalls et al. 2014 | rs6430538 | ACMSD/TMEM163 | 0.875 | 1.022 | 0.811 | FALSE |
| Nalls et al. 2014 | rs1474055 | STK39 | 1.214 | 1.156 | 0.127 | TRUE |
| Nalls et al. 2014 | rs115185635 | KRT8P25/APOOP2 | 1.142 | 1.284 | 0.355 | TRUE |
| Nalls et al. 2014 | rs12637471 | MCCC1 | 0.842 | 0.913 | 0.323 | TRUE |
| Nalls et al. 2014 | rs34311866 | TMEM175/GAK/DGKQ | 0.786 | 0.956 | 0.748 | TRUE |
| Nalls et al. 2014 | rs11724635 | BST1 | 1.126 | 1.129 | 0.155 | TRUE |
| Nalls et al. 2014 | rs6812193 | FAM47E/SCARB2 | 0.907 | 1.145 | 0.188 | FALSE |
| Nalls et al. 2014 | rs356182 | SNCA | 0.760 | 0.631 | 2.48E-08 | TRUE |
| Nalls et al. 2014 | rs9275326 | HLA-DQB1 | 0.826 | 0.748 | 0.006 | TRUE |
| Nalls et al. 2014 | rs199347 | GPNMB | 1.110 | 1.067 | 0.480 | TRUE |
| Nalls et al. 2014 | rs117896735 | INPP5F | 1.624 | NA | NA | NA |
| Nalls et al. 2014 | rs3793947 | DLG2 | 0.929 | 0.972 | 0.744 | TRUE |
| Nalls et al. 2014 | rs329648 | MIR4697 | 1.105 | 1.141 | 0.111 | TRUE |
| Nalls et al. 2014 | rs76904798 | LRRK2 | 1.155 | 1.025 | 0.811 | TRUE |
| Nalls et al. 2014 | rs11060180 | CCDC62 | 1.105 | 1.124 | 0.157 | TRUE |
| Nalls et al. 2014 | rs11158026 | GCH1 | 0.904 | 0.998 | 0.984 | TRUE |
| Nalls et al. 2014 | rs1555399 | TMEM229B | 0.897 | 0.999 | 0.991 | TRUE |
| Nalls et al. 2014 | rs2414739 | VPS13C | 1.113 | 1.239 | 0.028 | TRUE |
| Nalls et al. 2014 | rs14235 | BCKDK/STX1B | 1.103 | 1.228 | 0.012 | TRUE |
| Nalls et al. 2014 | rs17649553 | MAPT | 0.769 | 0.662 | 0.001 | TRUE |
| Nalls et al. 2014 | rs12456492 | RIT2 | 0.904 | 1.001 | 0.993 | FALSE |
| Nalls et al. 2014 | rs62120679 | SPPL2B | 1.097 | 1.035 | 0.682 | TRUE |
| Nalls et al. 2014 | rs8118008 | DDRGK1 | 1.111 | 1.066 | 0.478 | TRUE |
| Chang et al. 2017 | rs4653767 | ITPKB | 0.920 | 0.960 | 0.660 | TRUE |
| Chang et al. 2017 | rs34043159 | IL1R2 | 1.080 | 1.170 | 0.100 | TRUE |
| Chang et al. 2017 | rs353116 | SCN3A | 0.940 | 1.124 | 0.156 | FALSE |
| Chang et al. 2017 | rs4073221 | SATB1 | 1.100 | 0.999 | 0.996 | FALSE |
| Chang et al. 2017 | rs12497850 | NCKIPSD,CDC71 | 0.930 | 0.893 | 0.281 | TRUE |
| Chang et al. 2017 | rs143918452 | ALAS1,TLR9,DNAH1,BAP1,PHF7, NISCH,STAB1,ITIH3,ITIH4 | 0.680 | NA | NA | NA |
| Chang et al. 2017 | rs78738012 | ANK2,CAMK2D | 1.130 | 1.262 | 0.213 | TRUE |
| Chang et al. 2017 | rs2694528 | ELOVL7 | 1.150 | 1.341 | 0.025 | TRUE |

| | | | | | | |
|---|---|---|---|---|---|---|
| Chang et al. 2017 | rs9468199 | ZNF184 | 1.110 | 1.219 | 0.041 | TRUE |
| Chang et al. 2017 | rs2740594c | CTSB | 1.090 | 1.001 | 0.992 | TRUE |
| Chang et al. 2017 | rs2280104 | SORBS3,PDLIM2,C8orf58,BIN3 | 1.070 | 1.239 | 0.025 | TRUE |
| Chang et al. 2017 | rs13294100 | SH3GL2 | 0.920 | 0.984 | 0.854 | TRUE |
| Chang et al. 2017 | rs10906923 | FAM171A1 | 0.930 | 0.834 | 0.039 | TRUE |
| Chang et al. 2017 | rs8005172 | GALC | 1.080 | 1.162 | 0.070 | TRUE |
| Chang et al. 2017 | rs11343 | COQ7 | 1.070 | NA | NA | NA |
| Chang et al. 2017 | rs4784227 | TOX3 | 1.090 | 1.113 | 0.201 | TRUE |
| Chang et al. 2017 | rs601999 | ATP6V0A1,PSMC3IP,TUBG2 | 0.930 | 1.000 | 1.000 | TRUE |
| Foo et al. 2020 | rs246814 | SV3C | 1.11/1.07 | 1.053 | 0.683 | TRUE |
| Foo et al. 2020 | rs9638616 | WBSCR17 | 1.14/1.02 | 1.117 | 0.212 | TRUE |

Look-up of additional PD GWAS loci. STUDY, the original source of the data, SNP, the rs ID, GENE(s), the nearest genes. OR_STUDY, the reported odds ratio. OR_LARGE, the odds ratio in LARGE-PD. P_LARGE, the p-value from the LARGE-PD GWAS. CONCORDANCE indicates an agreement in effect size direction. Variants with an NA had a MAF less than 0.01 in LARGE-PD.

#### Supplementary Table 7: Admixture mapping fine-mapping results

| CHR | PEAK | GENE | TOP SNP | POS | PVAL | P_ADJ | MAF |
|---|---|---|---|---|---|---|---|
| 6 | 166465311-166607482 | RPS6KA2 | rs75880521 | 166524210 | 6.05E-04 | 0.352 | AFR:0.22; EUR: 0.0; AMR: 0.014 |
| 14 | 24713480-25147976 | STXBP6 | rs79647551 | 24719286 | 4.49E-05 | 0.070 | AFR:0.007; EUR:0.11; AMR:0.31 |
| 17 | 34404030-34456968 | NA | rs4795926 | 34449696 | 1.74E-02 | 1.000 | AFR:0.09; EUR:0.22; AMR:0.412 |
| 21 | 44767470-44915665 | ITGB2 | rs183517 | 44892765 | 1.66E-04 | 0.111 | AFR:0.22; EUR: 0.0; AMR: 0.014 |

LARGE-PD GWAS results overlayed with the admixture mapping peaks. CHR, the chromosomal location of the peak. PEAK, the position in hg19 coordinates. GENE, the gene co-localized with the peak. TOP SNP, the variant with the lowest p-value within the AM peak. POS, the variant position in hg19 coordinates. PVAL, the p-value from the LARGE-PD GWAS for the top variant. P_ADJ, the p-value adjusted for the number of imputed variants in the peak. MAF, the super-population frequencies of the variant from the 1000 Genomes Project.

**Supplementary Figure 1: Reference Samples for Ancestry analysis**

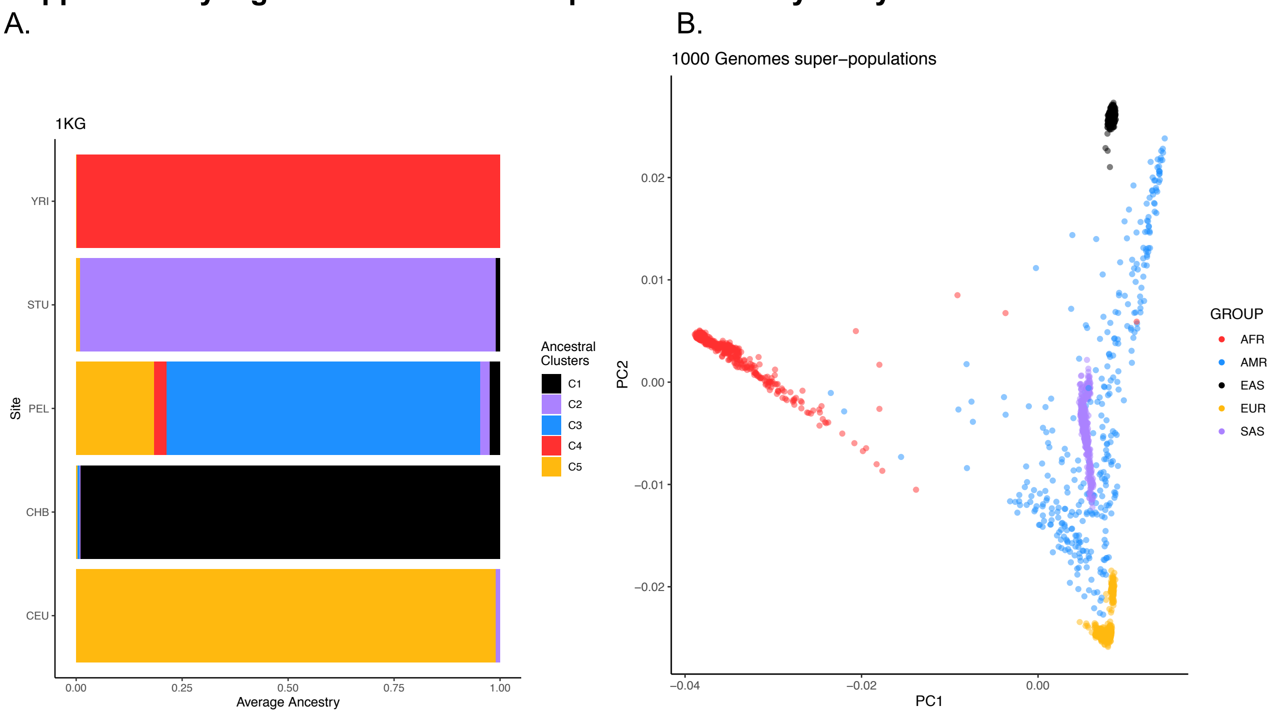


A: Mean ancestry proportions 1000 Genomes Project[74] populations used as references as determined by ADMIXTURE in an unsupervised manner. B: PCA plot of 1000 Genomes subjects used as references.

**Supplementary Figure 2: QQ plot of GWAS P-values**

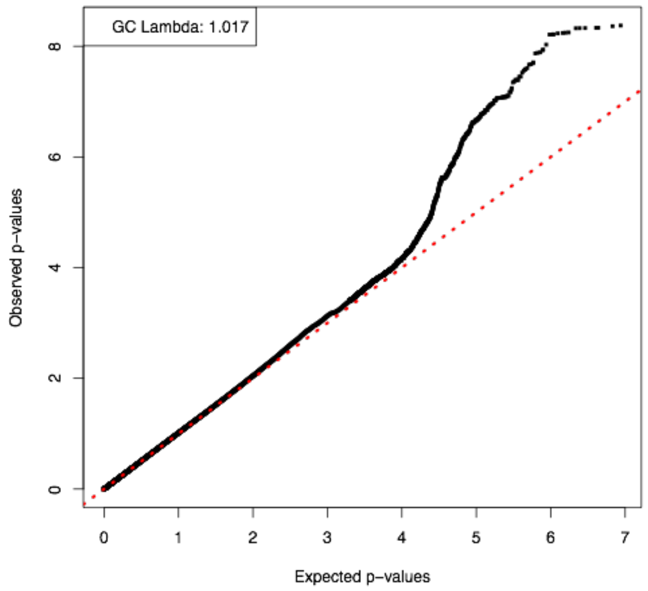


QQ plot of log-transformed p-values from the GWAS of imputed LARGE-PD data obtained via a logistic mixed model as implemented by the GENESIS package in R (see methods). Minimal inflation was observed; the genomic control lambda was 1.017.

**Supplementary Figure 3: Locuszoom-style plots of *SNCA* locus**

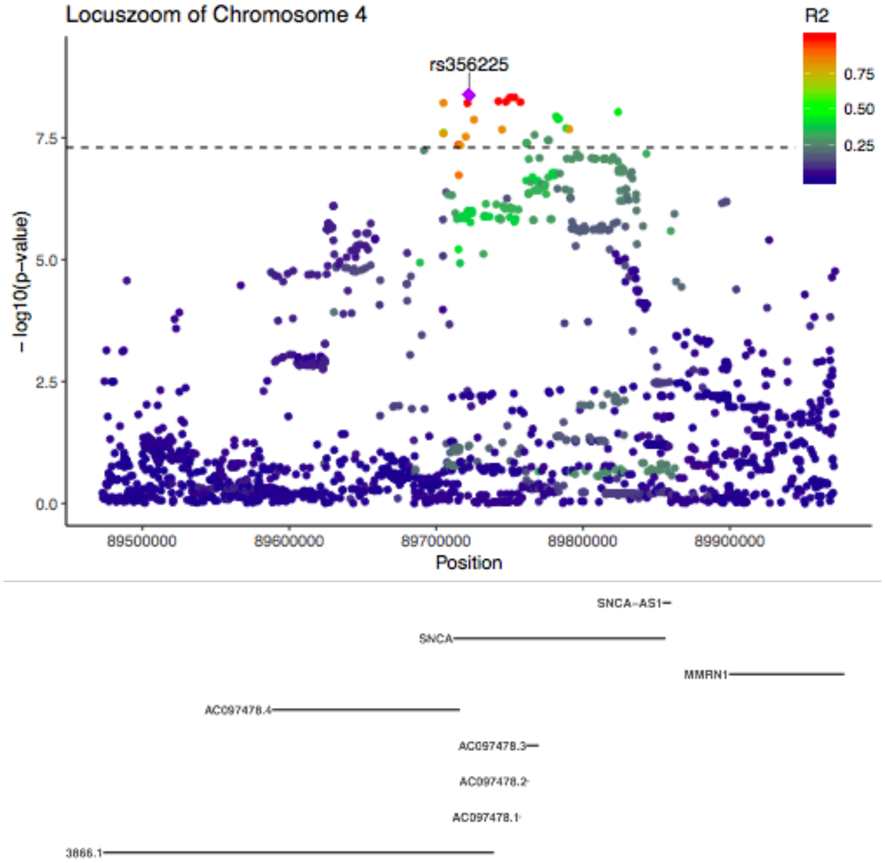


Locuszoom-style plots for the SNCA locus on chromosome 4. The plots generated via the Locuszoom tool are reliant on external LD data and might not be reflective of the true LD structure present in the GWAS dataset. Here, we use LD information estimated with PLINK to generate Locuszoom-style plots.

### Supplementary Figure 4: LD structure of genome-wide significant *SNCA* variants

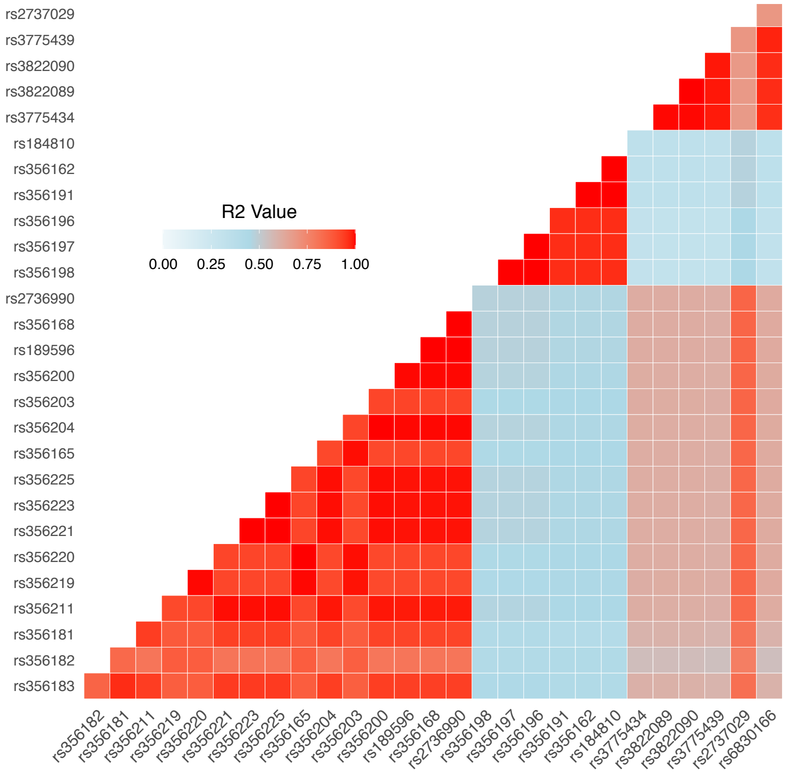


Linkage disequilibrium structure of genome-wide significant *SNCA* variants in the Peruvian subset of LARGE-PD. LD is estimated using PLINK and plotted in R. Note that 14 variants have a squared correlation with rs356182 greater than 0.8.

### Supplementary Figure 5: Conditional analysis of GWAS-significant *SNCA* variants

A. Conditioning on rs356182

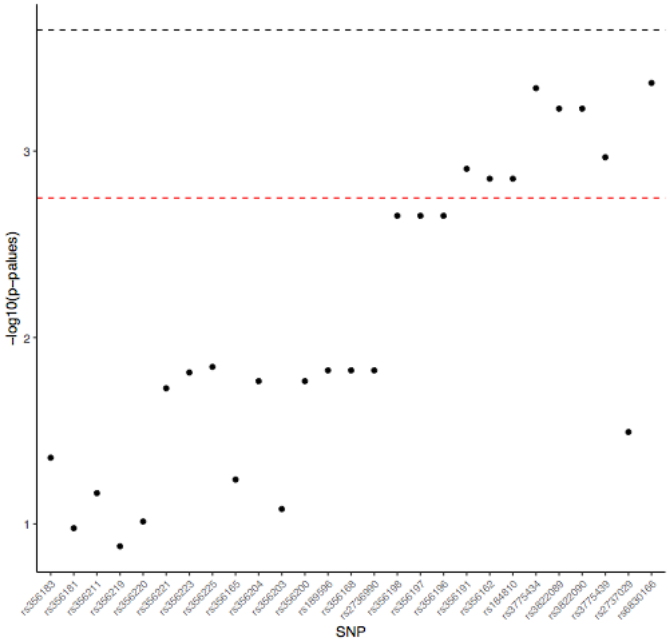

B. Conditioning on rs356182 and rs6830166

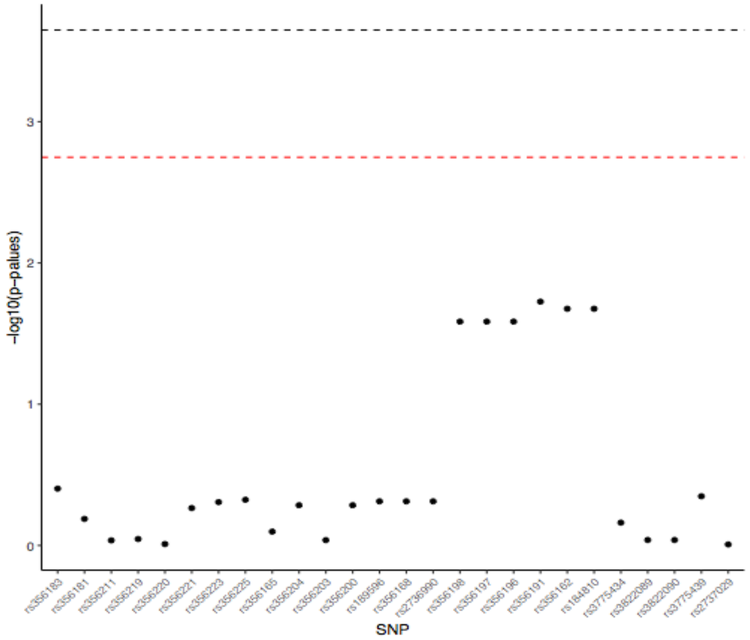


We first conditioned on rs356182, the lead variant from European-ancestry PD meta-analyses, followed by adding rs6830166, the SNP with the lowest P-value after conditioning on rs356182. We fit a logistic mixed model using the GMMAT in R, with the same covariates as our GWAS plus the SNP(s). The red dashed line displays a significance level adjusting for number of GWAS-significant variants; the black dashed line represents a regional significance level adjusting for 220 tests. Results are shown after conditioning on rs356182 (**A**) and rs356182 and rs6830166(**B**). Note that the evidence for independence depends on the chosen significance level.

### Supplementary Figure 6: Difference in study beta coefficients by MAF

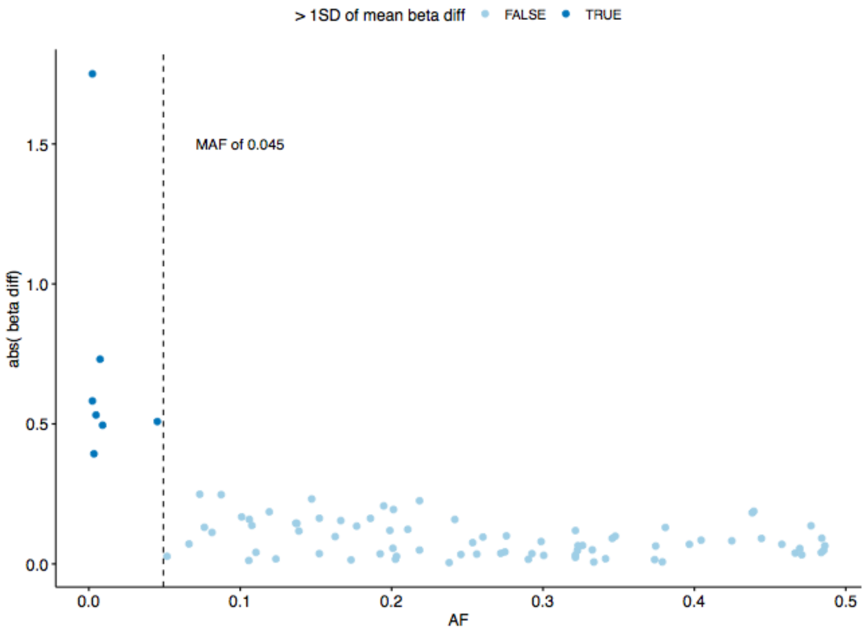


Six variants demonstrated a difference in beta coefficients from LARGE-PD and Nalls et al.[1] greater than one standard deviation of the mean. All of the variants have a MAF lower than 0.0452; three have a MAC lower than 10. This was partially due to the larger effect sizes of rare variants, but also could be attributed to inaccurate beta estimates due to insufficient sample size.

### Supplementary Figure 7: PD risk by ancestry

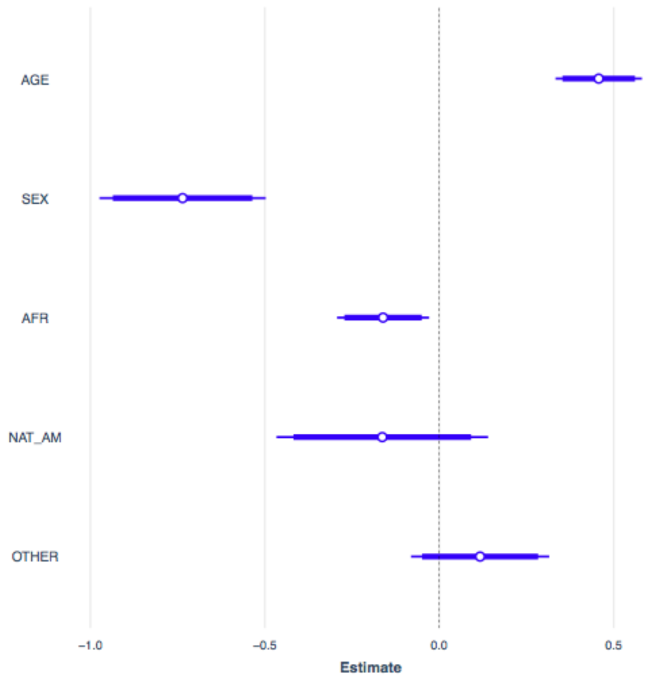


PD risk by ancestry as determined via logistic regression in R. Ancestry proportions were estimated using ADMIXTURE and a K of 5; two of the ancestry components that were not well-represented in

LARGE-PD are aggregated as “Other” in this analysis. The logistic model was fit adjusting for age, sex, LARGE-PD site, African ancestry proportion, East Asian ancestry proportion, and Amerindian ancestry proportion; we limited the analysis to the unrelated subset of LARGE-PD. The European ancestry proportion is used as the reference. Age, sex, and African ancestry were significant (P-value < 0.05). Beta coefficients were plotted using the jtools package in R.

#### Supplementary Figure 8: Admixture mapping single-ancestry models

A. African

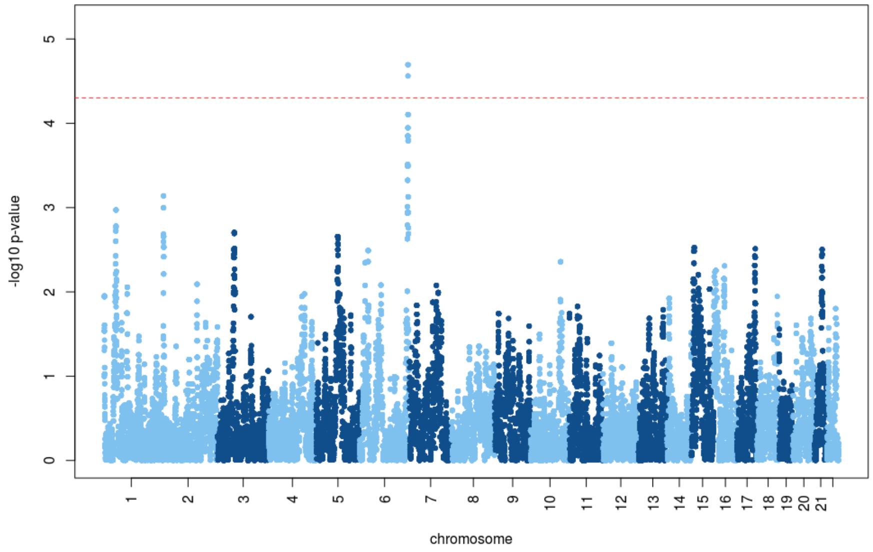


B. Native American

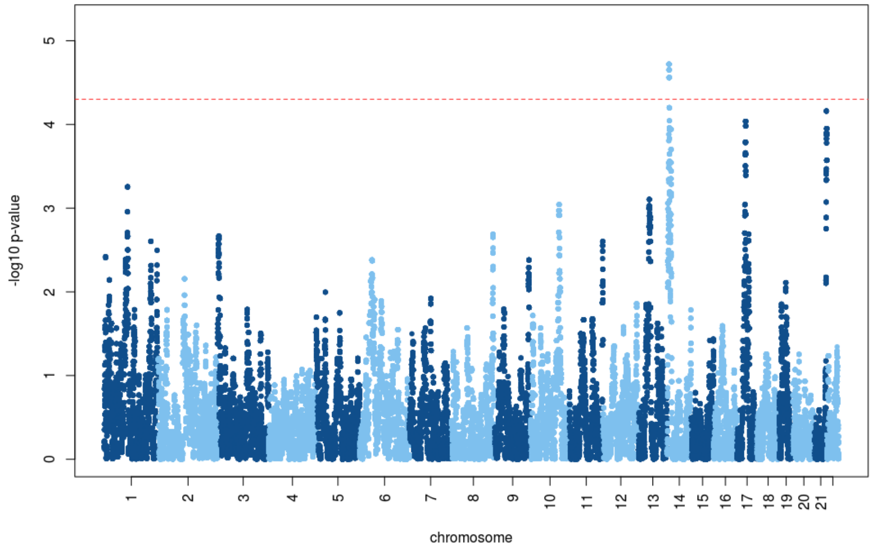


C. European

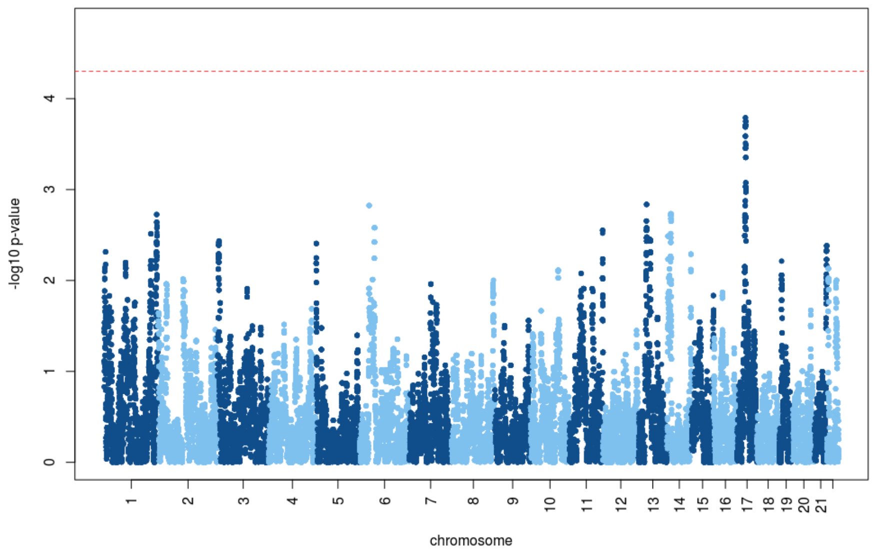


A: Results from admixture mapping as implemented in GENESIS including only the African local ancestry term. Dashed red line indicates statistical significance (**see supplementary methods**)

B: Results from admixture mapping as implemented in GENESIS including only the Amerindian local ancestry term. Dashed red line indicates statistical significance (**see supplementary methods**)

C. Results from admixture mapping as implemented in GENESIS including only the European local ancestry term. Dashed red line indicates statistical significance (**see supplementary methods**)
